# Nature prescriptions: a scoping review with a nested meta-analysis

**DOI:** 10.1101/2022.03.23.22272674

**Authors:** Phi-Yen Nguyen, Hania Rahimi-Ardabili, Xiaoqi Feng, Thomas Astell-Burt

**Affiliations:** Population Wellbeing and Environment Research Lab (PowerLab), School of Population Health, University of New South Wales, Sydney, New South Wales, Australia; School of Public Health and Preventive Medicine, Monash University, Melbourne, Victoria, Australia; Centre for Health Informatics, Australian Institute of Health Innovation, Macquarie University, Sydney, New South Wales, Australia; Population Wellbeing and Environment Research Lab (PowerLab), School of Health and Society, Faculty of Arts, Social Sciences, and Humanities, University of Wollongong, Wollongong, New South Wales, Australia

## Abstract

**Background:** “Nature prescriptions” are gaining popularity as a form of social prescribing and in response to calls for sustainable healthcare. Our review and meta-analysis appraised evidence of effectiveness of nature prescriptions on various health outcomes. In doing so, we sought to determine the factors that are critical for the success of nature prescriptions, based on Social Cognitive Theory.

**Methods:** This is a scoping review with a nested meta-analysis for a subset of outcomes. Five databases were searched up to July 25, 2021. Randomised and non-randomised controlled studies featuring a nature prescription (i.e. an instruction or organised programme, by a health or social provider, to promote spending time in nature) are included. All health outcomes are eligible, but only key pre-specified outcomes are qualified for meta-analysis. Two reviewers independently conducted all steps of study selection; one reviewer conducted data collection and risk of bias assessment. Summary data was extracted from published reports for analysis. Random-effect models for meta-analysis were conducted using Review Manager 5.4.1.

**Findings:** We identified 86 unique studies (116 reports), of which 26 studies contributed data to meta-analysis. Compared to control, nature prescription programmes resulted in a greater reduction in systolic blood pressure (MD = -4·9mmHg [-9·6 to -0·1], I^2^=65%) and diastolic blood pressure (MD = -3·6mmHg [-7·4 to 0·1], I^2^=67%). They also had a moderate-to-large effect on depression scores (SMD=0·5 [0·2 to 0·8], I^2^=79%) and anxiety score (SMD=0·6 [0·1 to 1·2], I^2^=90%). Lastly, they resulted in a greater increase in daily step counts (MD = 900 steps [790-1010], I^2^=0%), but did not improve weekly time of moderate physical activities (MD = 25·9 minutes [-10·3 to 62·1], I^2^=53%). Most studies have moderate to high risk of bias, principally due to non-blinding nature of the interventions, small sample size and lack of analysis plan to rule out risks of bias.

**Interpretation:** Nature prescription programmes may provide cardiometabolic and mental health benefits and increase physical activity. Effective nature prescription programmes can select from a range of natural settings, activities and might be implemented via social and community channels, besides health providers. The Social Cognition Theory is useful in designing future nature prescription programmes.

**Funding:** This work was supported by the Hort Frontiers Green Cities Fund, part of the Hort Frontiers strategic partnership initiative developed by Hort Innovation, with co-investment from the University of Wollongong (UOW) Faculty of Social Sciences, the UOW Global Challenges initiative and contributions from the Australian Government (project number #GC15005). T.A-B. was supported by a National Health and Medical Research Council Boosting Dementia Research Leader Fellowship (#1140317). X.F. was supported by a National Health and Medical Research Council Career Development Fellowship (#1148792).

**Panel: Research in context**

Evidence before this study
Extensive evidence indicates contact with nature is associated with social, mental and physical health. However, little evidence exists on the effectiveness of nature prescriptions, which involve a health provider (e.g. general practitioner) recommending a patient to spend a fixed amount of time a week in a natural setting (e.g. a park). Other studies have attempted to evaluate the benefits of food prescription or green prescription programmes, which do not necessarily involve nature exposure. Only one systematic review on nature prescriptions has been conducted to date, which is a qualitative review without meta-analysis. The review concluded that the evidence (studies up to June 2019) was too sparse to discern any clear evidence of health impacts. There was insufficient information to assess the risk of bias or quality of evidence in the review. Moreover, the review included only nature prescriptions dispensed in outpatient settings, which left out prescription programmes implemented by other institutions, such as welfare centres, social services, universities or workplaces.

Added value of this study
Our review is the first to provide comprehensive appraisal including meta-analysis of the effectiveness of nature prescription programs on multiple health outcomes. The scoping review identified a range of promising nature-based interventions that were dispensed outside the clinic setting and did not self-label as a nature prescription, but would be effective as one. The nested meta-analyses on key outcomes demonstrated positive benefits on blood pressure, symptoms of depression and anxiety, and physical activity levels.

Implications of all the available evidence
Our findings suggest that an effective nature prescription programme can select from a range of natural settings, activities and can be implemented via social and community channels, in addition to health providers. In addition, we also demonstrated that the Social Cognition Theory framework is useful in designing future nature prescription programmes.

## 1. INTRODUCTION

Extensive evidence indicates contact with nature is associated with social, mental and physical health ^1–3^. These potential benefits include favourable pregnancy outcomes ^4^ through to reduced risks of cardiometabolic ^5,6^ and neurodegenerative diseases ^7,8^ in older adults. While addressing the well-documented inequities in green space ^9^ are warranted, improving provision will be insufficient to ensure everyone benefits ^10^.

Nature prescriptions have emerged as a potential solution to enable and empower people to spend more time in nature where that was not previously the case. Nature prescriptions are an adjunct to conventional healthcare, such as the educational and pharmaceutical treatment of non-communicable diseases. A nature prescription typically involves a health provider (e.g. general practitioner) recommending a patient to spend a fixed amount of time a week in a natural setting, such as a park. It is widely considered that the benefits of nature prescribing will reach far beyond health, such as increasing social connectedness ^11^ and pro-environmental behaviours ^12^.

To our knowledge, only one systematic review has been conducted on nature prescription to date. This qualitative review by Kondo and colleagues identified eleven nature prescription studies published up to June 2019 ^13^ and concluded that the evidence was too sparse to discern any clear evidence of health impacts. Another study adopted a survey approach to investigate the benefits of green space programmes for mental health ^14^. From 2020 onwards, we noted a substantial upswing in interest and publication of new nature prescription studies. This raises the potential for meta-analysis and critical appraisal of the importance of personal/cognitive, behavioural and environmental factors to the success of these intervention programmes using Social Cognitive Theory (SCT) ^15^.

Accordingly, the objective of this review is to identify evidence for effective nature prescriptions and to determine the factors which are critical for their success. We pose the following questions:

a. To what extent can nature prescriptions improve social, mental and physical health?
b. What are the design characteristics of nature prescriptions with demonstrated health benefits?
c. What are potential channels to dispense a nature prescription beside a clinic or hospital?

## 2. METHODS

Reporting of this review was guided by the Preferred Reporting Items for Systematic Reviews and Meta-Analyses (PRISMA) guidelines ^16^. This review was not registered *a priori*.

### 2.1. Search strategy

We searched the following databases for articles from inception up to July 25, 2021: MEDLINE via Ovid, Embase via Ovid, PsycINFO via Ovid, CINALH via EBSCO and CENTRAL via Cochrane Library. The search was supplemented by manual search of reference list from relevant systematic reviews. The search strategy is available in Supplementary file S1.

### 2.2. Study selection

Two reviewers (PN and HA) independently screened all titles and abstracts in duplicate and excluded studies that did not meet inclusion criteria (Table 1). Full texts of selected articles were reviewed by one reviewer (PN) and checked by a second reviewer (H-RA). Disagreement was resolved by discussion with senior reviewers (XF and TA-B). All stages of study screening were conducted using Covidence (Veritas Health Innovation, Australia). We excluded interventions with a dietary focus as these have been previously investigated ^17^.

**Table 1.** Eligibility criteria for study selection.

|  | **Inclusion** | **Exclusion** |
| --- | --- | --- |
| **Participant** | Any participant | Animal studies |
| **Intervention** | An instruction by a health or social provider to patients to spend time in a nature setting, such as a park; or<br>Any programme organised by a health or social institutions for their patients or clients that features nature-based interventions.<br>Nature-based interventions are defined as interventions that:<br>• Used nature-based therapy to improve health outcomes; and<br>• Involved exposure to a nature environment, including green spaces and blue spaces<br>Multimodal programmes where one component is nature-based activities are eligible. | 1) Interventions aimed at only changing the environment in which people live:<br>• Building new green spaces, changing design or providing facilities within green spaces<br>• Provision of gardens, indoor vegetation, community allotments, outdoor gyms etc. without organising any activity<br>2) Programmes requiring high levels of safety and skilled organisers (e.g. wilderness adventure programmes, animal-assisted therapies, mountain hiking, etc.)<br>3) Simulation of nature spaces (e.g. virtual reality, photos, audio records) without actual nature exposure<br>4) School and after-school curricular activities, or any interventions aimed at increasing play time without a clear nature focus |
| **Control** | No intervention or intervention taking place in a non-nature setting | No control group |
| **Outcomes** | Physical, psychological/cognitive health and behavioural outcomes | Studies that only measure these outcomes:<br>• Social, economic and financial outcomes<br>• Diet composition and dietary patterns |
| **Study design** | Randomised controlled trials (RCTs), non-randomised or quasi-randomised controlled trials | Observational studies<br>Qualitative studies<br>Conference abstracts/proceedings, editorials, thesis, letters to editors, short reports |
| **Other** | .. | Non-English studies |

### 2.3 Data collection

One reviewer (PN) extracted data using a standardised extraction form. Data extracted included characteristics of studies, participants, interventions and outcomes. Study characteristics included study design, sample size and location. Participant characteristics included social background, pre-existing medical conditions and age groups, as defined in the eligibility criteria. Interventions were characterised based on the nature setting where the intervention took place, types of activities undertaken by participants, whether the nature setting was indoor or outdoor, and the institutions who introduced the participants to the intervention (‘referring institutions’). The referring institutions must be have an established medical or social connection to the patients, such as treating hospitals, social services, welfare centres, etc. We recorded “None” if the participants were recruited through standard trial recruitment methods, such as mass emails, academic recruiters or public bulletins. We categorised the outcomes measured as physical, psychological/cognitive or behavioural outcomes. Biomarkers were recorded separately. We also recorded specific outcomes where a positive benefit was reported based on 95% confidence intervals or p-value <0·05 (if 95% confidence intervals were not available), and recorded whether the findings were based on within-group (pre-vs post-intervention) comparisons or between-group (intervention vs control group) comparisons.

We planned to conduct a nested systematic review and meta-analysis for the following outcomes: systolic blood pressure (SBP), diastolic blood pressure (DBP), depression, anxiety, step counts and time spent on physical activities. Therefore, for studies that reported these outcomes, we additionally recorded the means and standard deviations for both groups, either as changes from baseline or post-intervention measurements, whichever available. If not provided, standard deviations (SD) were calculated from standard errors or 95% confidence intervals of the mean ^18^. If an outcome was measured at multiple follow-ups, we selected the time point most often reported amongst all studies, to make results more comparable amongst studies. If an outcome was measured using multiple scales, we record the scale most often reported amongst all studies. In one study, metabolic equivalent of task (MET) minutes were converted to minutes spent doing moderate physical activities by dividing means and SD by a factor of four ^19^. Mean changes from baseline and post-intervention means were synthesised separately in subgroup meta-analyses, and their results were pooled together in the final meta-analysis. Studies that provided no extractable data or no data to calculate SD were excluded from meta-analysis and presented narratively.

### 2.4. Risk of bias assessment

Risk of bias assessment was conducted by one reviewer (PN) for studies included in the nested meta-analysis, using the ROBINS-I tool for non-randomised studies and the ROB 2.0 tool for randomised trials.

### 2.5. Statistical analysis

We performed descriptive statistics (frequency and percentage) of intervention characteristics, including participant age groups, settings and activity types, as well as the referring institutions.

We assume the true treatment effects would likely differ among studies due to heterogeneity in age groups, pre-existing health conditions and intervention characteristics. Hence, we used DerSimonian-Laird random-effect models for meta-analysis of all outcomes. Standardised mean differences were used in meta-analysis of depression and anxiety, which were measured using various scales, and interpreted based on rule of thumb (0.2 as small effect, 0.5 as moderate effect, 0.8 as large effect). For other outcomes, mean differences were used. If both mean changes from baseline and post-intervention means were reported, post-intervention means were used. All analyses were conducted in Review Manager 5.4.1.

## 3. RESULTS

We retrieved a total of 5,115 records from 5 databases, with an additional of 6 studies from backward/forward citation checking during screening. The final sample consisted of 86 unique studies (116 reports). The study selection process is summarised in Figure 1. The list of excluded full texts with reasons is in Supplementary File S3.

**Figure 1.**
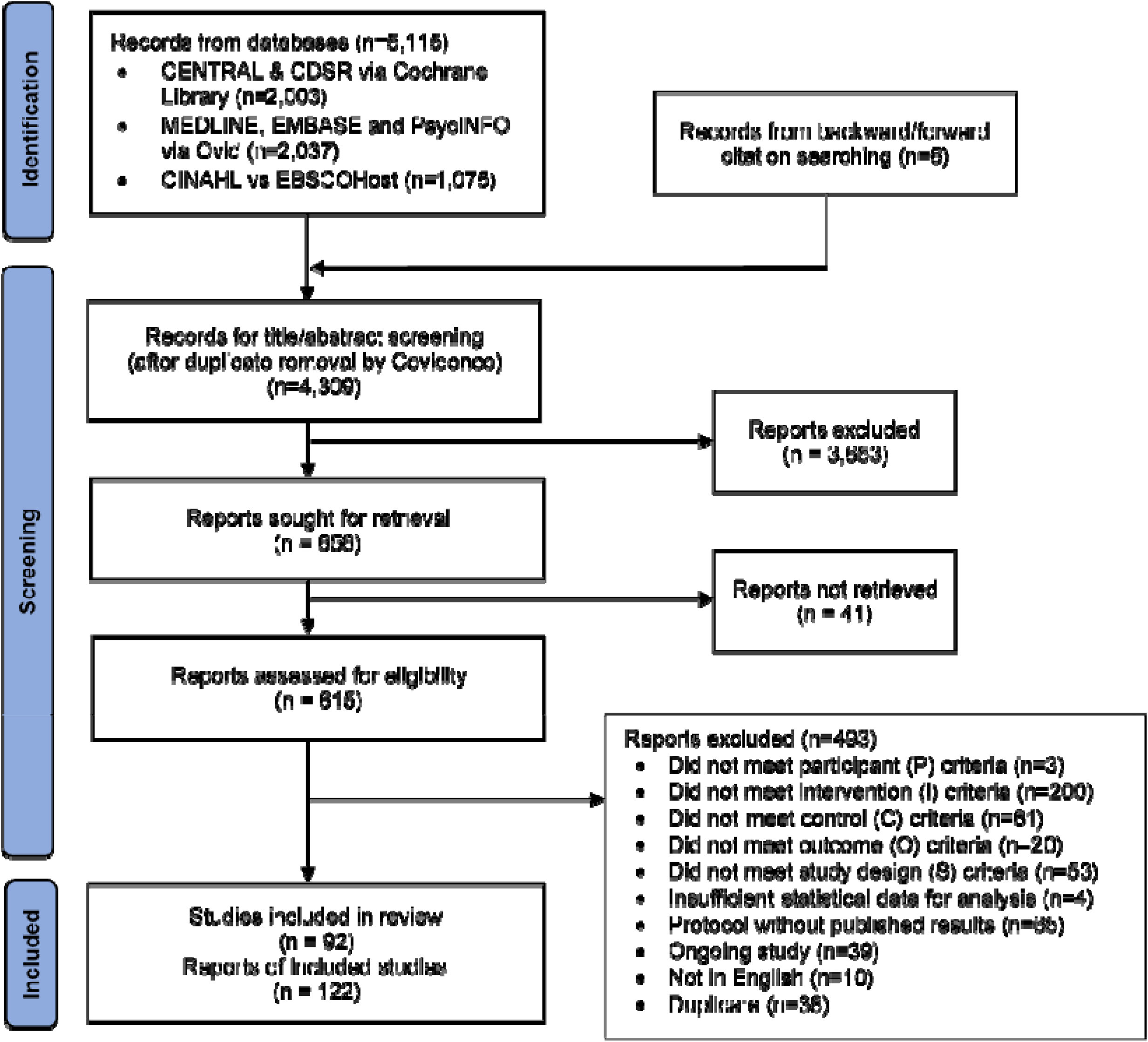
PRISMA flowchart of record retrieval and selection.

### 3.1. Study & participant characteristics

The included studies ranged from 1999 to 2021, with a significant drop in publication in 2019, possibly due to influences of the COVID-19 pandemic (Supplementary Figure S1). Most included studies are randomised controlled trials (n=67, 73%). Most studies are concentrated in high-income countries (Supplementary Figure S2). Countries where most interventions took place were South Korea (n=18, 20%), the United States (US) (n=16, 17%) and Japan (n=10, 11%). The studies examined a diverse range of age groups, mainly adults (n=59, 64%) or elderly (n=25, 27%). Only 11 studies (12%) involved participants under the age of 18. Eleven studies (12%) specifically recruited participants with socioeconomically-disadvantaged backgrounds, such as low-income families or minor ethnic groups. The most common pre-existing conditions are psychiatric disorders (e.g. schizophrenia, ADHD, etc.) (n=13, 14%), cardiovascular disorders (e.g. post-stroke, congestive heart failure, etc.) (n=12, 13%) and musculoskeletal disorders (fibromyalgia, history of falls or balancing issues, etc.) (n=6, 7%).

Risk-of-bias assessments are available in Supplementary File S2. The most important concerns for risk of bias were missing outcome data (due to high rates of dropouts without explicit reasons) and bias from measurement of outcomes (due to non-blinding nature of the intervention and the subjective nature of psychological assessment scales used).

### 3.2. Intervention characteristics

All included studies feature aspects of a nature prescription i.e. instructing the participants to spend time engaging with nature at various capacities. Only four studies, however, identified themselves as a nature or park prescription intervention ^20–23^. The most common settings for such nature-based therapy are forests and nature reserves (n=32, 35%), parks (n=26, 28%), small community or home gardens (n=15, 16%), or botanical gardens/allotments (n=10, 11%). Two studies (2%) also featured blue spaces such as beaches. The most common activities recommended to participants were walking in nature (n=42, 46%), farming or gardening (n=27, 29%) and mindfulness exercises (e.g. meditation, breathing exercises) (n=27, 29%), among a range of other activities (e.g. art and craft, group sports, reading or listening to music, etc.). Seven studies (8%) allowed participants to freely choose their activities ^20–26^.

Participants were commonly introduced to the trials by their health providers (n=23, 24%) or community service providers (n=24, 26%). The community service providers were diverse in nature, and tended to be associated with pre-existing conditions of the participants (e.g. day care services or senior centres for elderly in long-term care, job rehabilitation centres for people on extended sick leave or welfare centres for low-income families). The health providers were also varied, ranging from general practitioner (GP) clinics, family health centres, post-stroke rehabilitation centres to hospitals.

We evaluated the design of all interventions to see if they demonstrate aspects of the Social Cognitive Theory (SCT) framework for behavioural change i.e. an increased engagement with nature. All studies featured behavioural factors such as selecting activities that participants can easily carry out on their own (n=61, 66%), or providing training (n=46, 50%) or tools (n=37, 40%) to assist with the activities e.g. gardening equipment, exercise equipment or maps of walking paths. Most studies (n=77, 84%) featured environmental factors such as conducting activities in group for peer support (n=52, 57%), selecting nature sites within the proximity of participants’ home, their regular health providers’ or community service providers’ offices (n=38, 41%). In twelve studies (13%), the authors mentioned providing measures to enable access such as transportation or free tickets for gym entry. However, the third aspect of the SCT, cognitive factors, were only featured in a third of studies (n=26, 28%) such as educating participants on the benefits of nature exposure (n=18, 20%) and setting goals to motivate participants (n=17, 18%).

Table 2 provides summary statistics for all included studies. Intervention characteristics and evaluation of programmes based on SCT framework are available in Supplementary File S4.

**Table 2.** Study characteristics.

|  | Frequency (%) |
| --- | --- |
| **Study characteristics** |  |
| Study design |  |
| Randomised controlled studies | 67 (73%) |
| Non-randomised controlled studies | 25 (27%) |
| Study location |  |
| South Korea | 18 (20%) |
| United States of America | 16 (17%) |
| Japan | 10 (11%) |
| United Kingdom | 7 (8%) |
| China | 5 (5%) |
| Other | 36 (39%) |
| **Participant characteristics** |  |
| Age group |  |
| Children (<10 years old) | 9 (10%) |
| Adolescent (10-18 years old) | 2 (2%) |
| Adults (18-65 years old) | 59 (64%) |
| Elderly (>65 years old) | 25 (27%) |
| Social background of participants |  |
| University students | 16 (17%) |
| Socioeconomically-disadvantaged background | 11 (12%) |
| Military members | 2 (2%) |
| Office workers | 2 (2%) |
| Long-term care residents | 2 (2%) |
| Underlying health conditions |  |
| Psychiatric disorders | 13 (14%) |
| Cardiovascular disorders | 12 (13%) |
| Musculoskeletal disorders | 6 (7%) |
| Cancer | 4 (4%) |
| Neurological disorders | 4 (4%) |
| Sexual health | 2 (2%) |
| Respiratory disorders | 1 (1%) |
| Substance use disorder | 1 (1%) |
| **Intervention characteristics** |  |
| Identified as 'nature' or 'green prescription' | 4 (4%) |
| Setting of nature-based therapy |  |
| Forests & nature reserves | 32 (35%) |
| Parks | 26 (28%) |
| Small gardens | 15 (16%) |
| Botanical gardens | 10 (11%) |
| Farms | 5 (5%) |
| Other urban green spaces | 5 (5%) |
| Greenhouses | 2 (2%) |
| Beaches | 2 (2%) |
| Activities taken by participants |  |
| Walking | 42 (46%) |
| Farming/gardening | 27 (29%) |
| Mindfulness & relaxation | 25 (27%) |
| Other physical exercises | 23 (25%) |
| Group games, including sports | 7 (8%) |
| Art & craft | 4 (4%) |
| Socialising activities, including dance | 3 (3%) |
| Enjoying nature & relaxation | 2 (2%) |
| Listening to music | 1 (1%) |
| Any activity chosen by participants | 7 (8%) |
| Institutions introducing participants to intervention |  |
| Health providers | 23 (25%) |
| Welfare & community service providers | 24 (26%) |
| Employers | 4 (4%) |
| Probation service centre | 2 (2%) |
| Long-term care providers | 2 (2%) |
| Schools | 1 (1%) |
| **Application of Social Cognitive Theory framework** |  |
| **Behavioural factors** | **92 (100%)** |
| Participants able to carry out activities on their own | 61 (66%) |
| Training provided | 46 (50%) |
| Tools provided | 37 (40%) |
| **Environmental factors** | **77 (84%)** |
| Activities conducted in groups | 52 (57%) |
| Nature sites accessible to participants | 38 (41%) |
| If not, measures in place to enable access | 12 (13%) |
| **Cognitive factors** | **26 (28%)** |
| Informing participants on benefits of nature exposure | 18 (20%) |
| Setting goals for participants | 17 (18%) |
| **Evidence of health benefits** |  |
| Physical health outcomes | 39 (42%) |
| Psychological & cognitive outcomes | 62 (67%) |
| Behavioural outcomes | 23 (25%) |
| Biomarkers measured | 20 (22%) |

### 3.3. Health outcomes

Thirty-nine studies (42%) reported benefits on outcomes related to physical health. Outcomes measured tended to be specific to the pre-existing health conditions. For example, interventions addressing cardiovascular disorders reported benefits on cardiometabolic indicators such as blood pressures, heart rates, aerobic fitness and body weight. Interventions for musculoskeletal and neurological disorders reported benefits on pain and various gross motor function tests such as Timed Sit-to-Stand or Timed Up-and-Go.

Two-third of studies (n=62, 67%) reported benefits on psychological or cognitive outcomes. A diverse range of measurement scales were used, mainly to assess moods (e.g. Profile of Mood States), depression (e.g. Beck’s Depression Inventory), stress (e.g. Perceived Stress Scale), anxiety (e.g. State-Trait Anxiety Scale) and quality of life (e.g. 36-item Short-Form Survey).

Twenty-three studies (25%) reported improved behavioural outcomes, mainly time spent outdoor, time spent on moderate-vigorous physical activities and step counts via pedometers. Eleven of these studies (58%) featured all three components of the SCT framework.

Twenty studies (22%) measured various biomarkers, mainly indicators of stress (e.g. salivary cortisol) and inflammatory responses (e.g. cytokines) and components of the haemodynamic control system (e.g. endothelin-1, AT1 receptors).

Table 3 provides a summary of findings for all included studies.

**Table 3:**
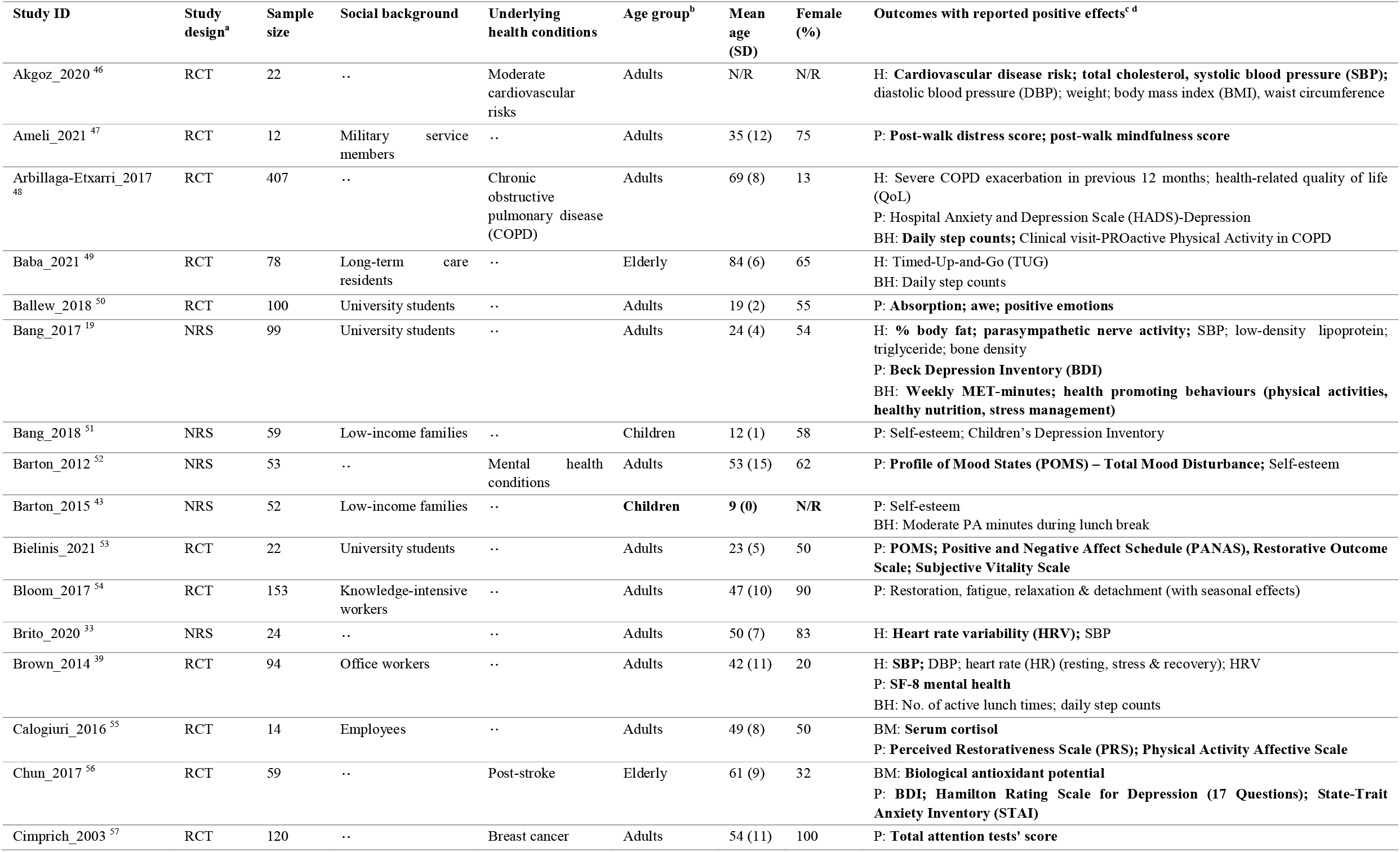
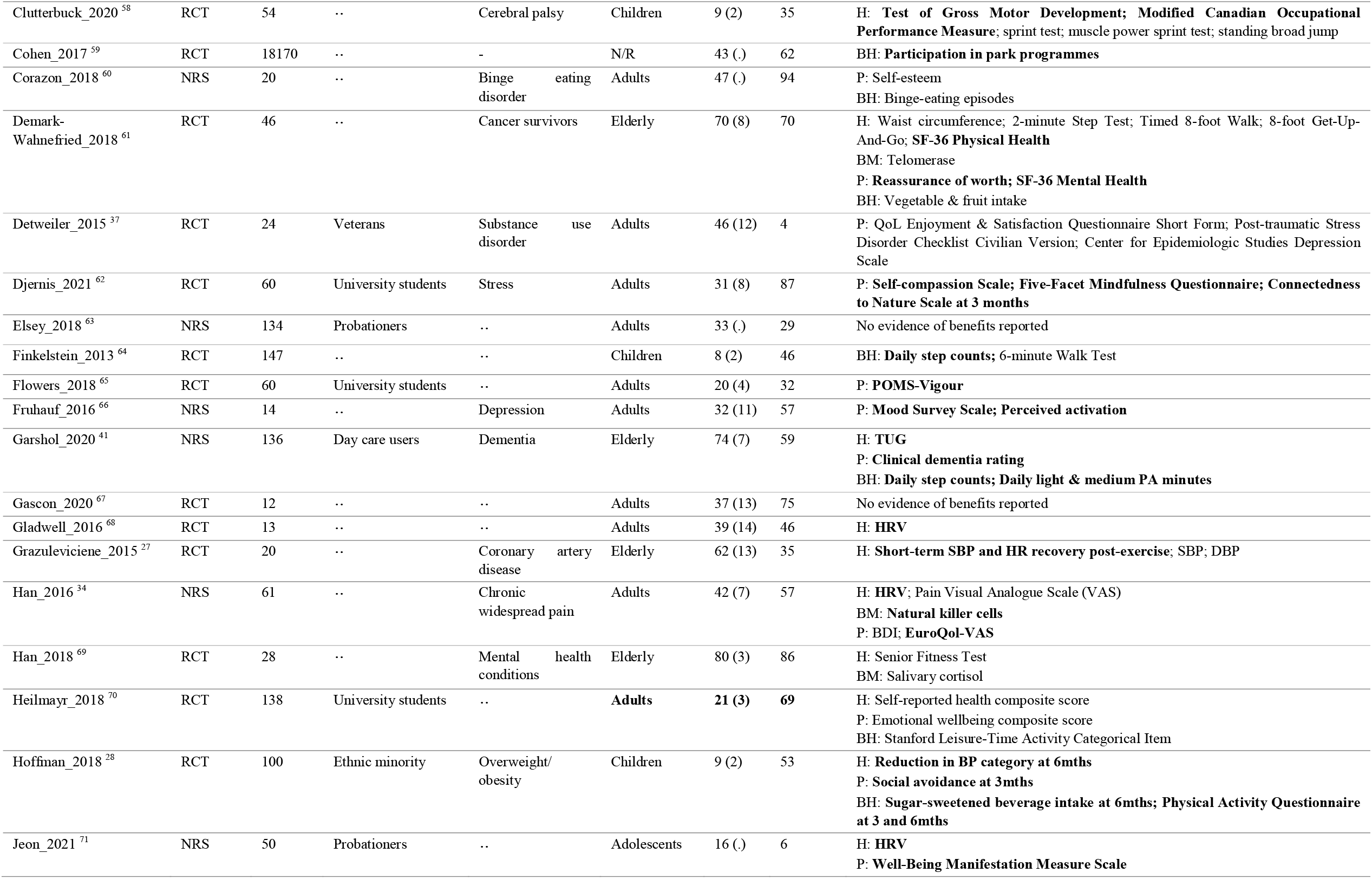
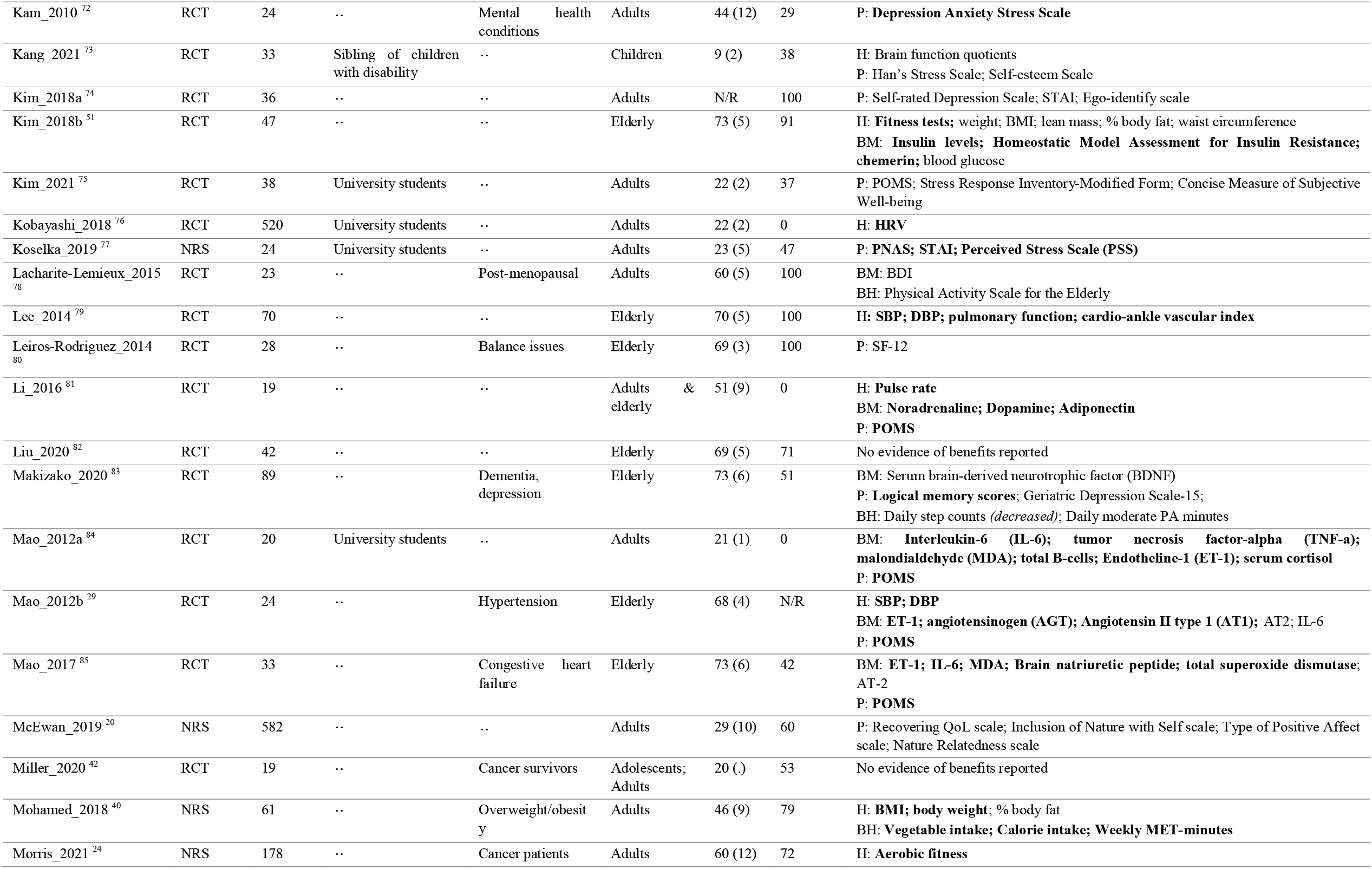
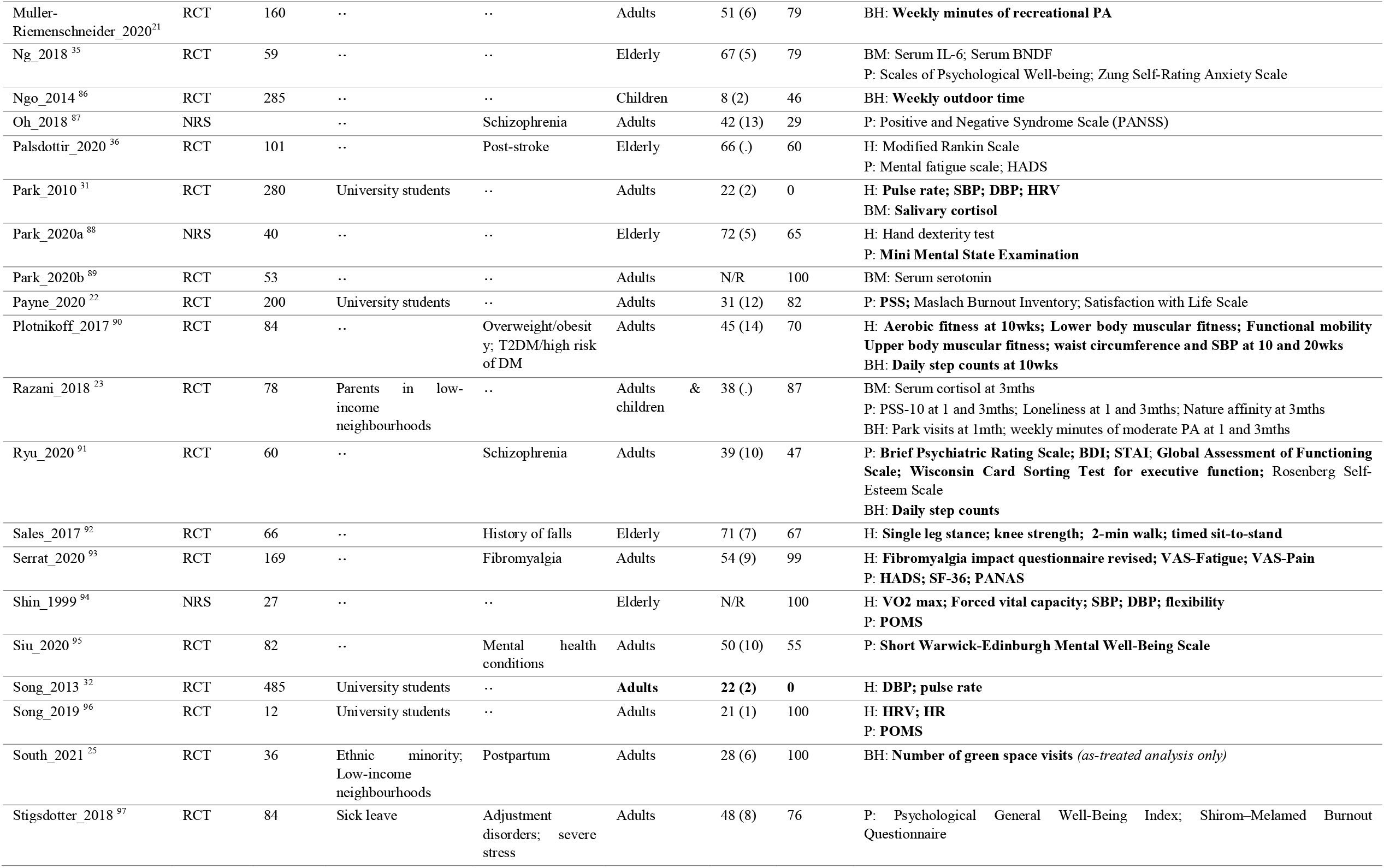
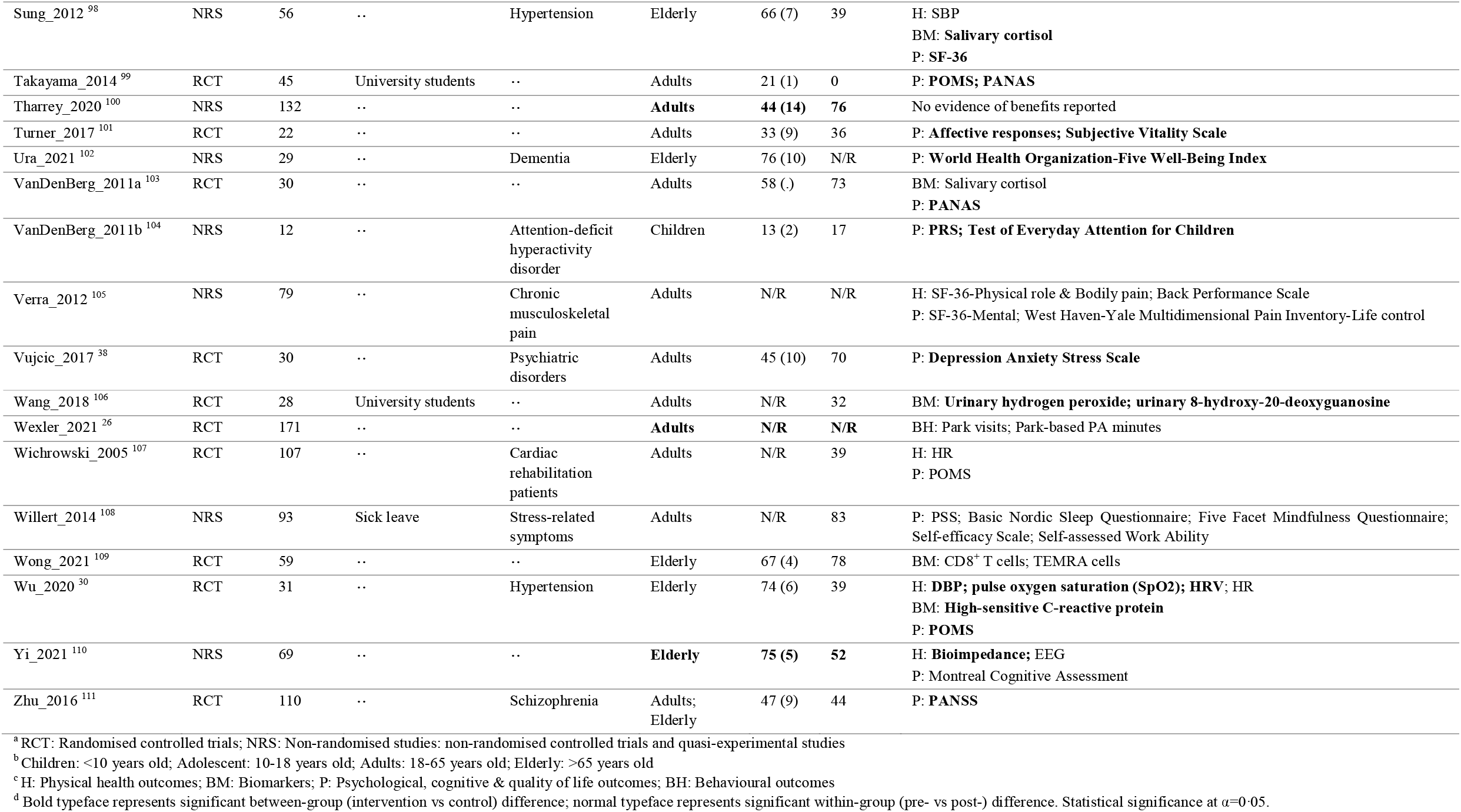
Summary of study designs and findings.

#### 3.3.1. Blood pressures

Five RCTs and two non-randomised studies (NRSs) contributed data to the meta-analysis (Figures 2A-B). The follow-up time ranged from 1 week to 12 weeks from baseline, except for one study ^27^ which conducted baseline and follow-up measurements within the same day. Compared to control, nature prescription programmes resulted in a greater reduction in SBP (MD = -4·9mmHg, 95% CI -9·6 to -0·1, I^2^=65%) and DBP (MD = -3·6mmHg, 95% CI -7·4 to 0·1, I^2^=67%).

**Figure 2.**
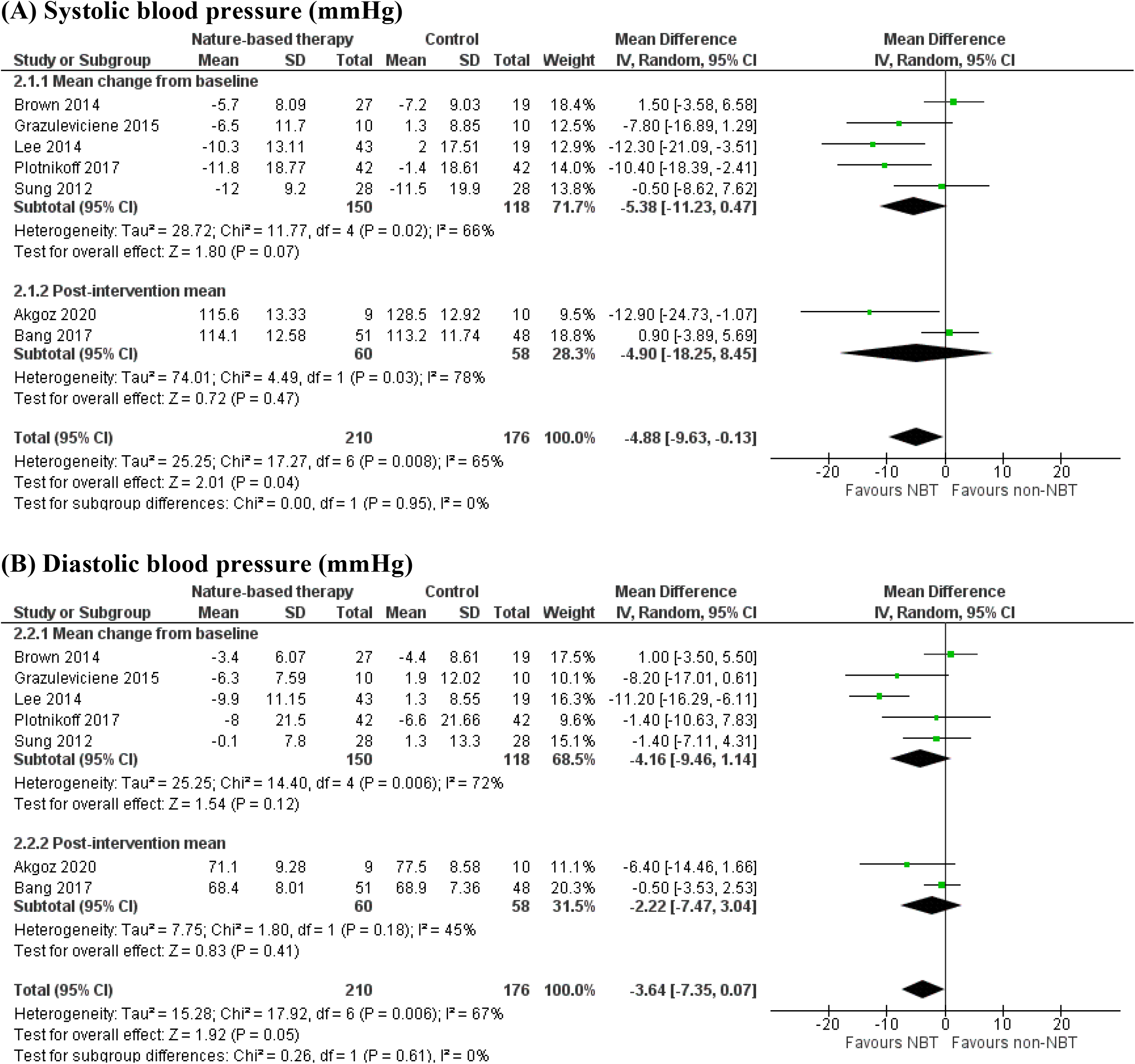
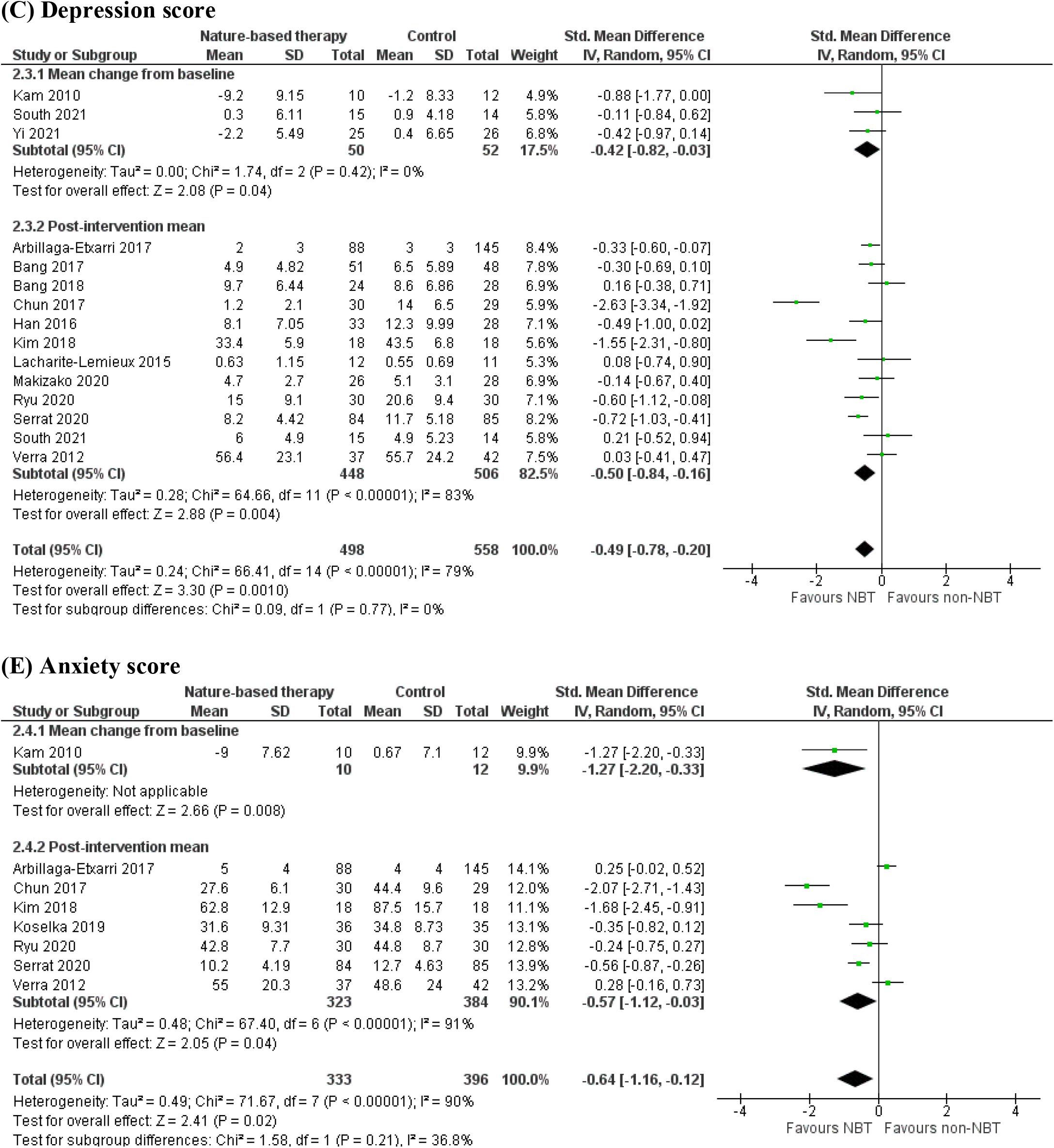
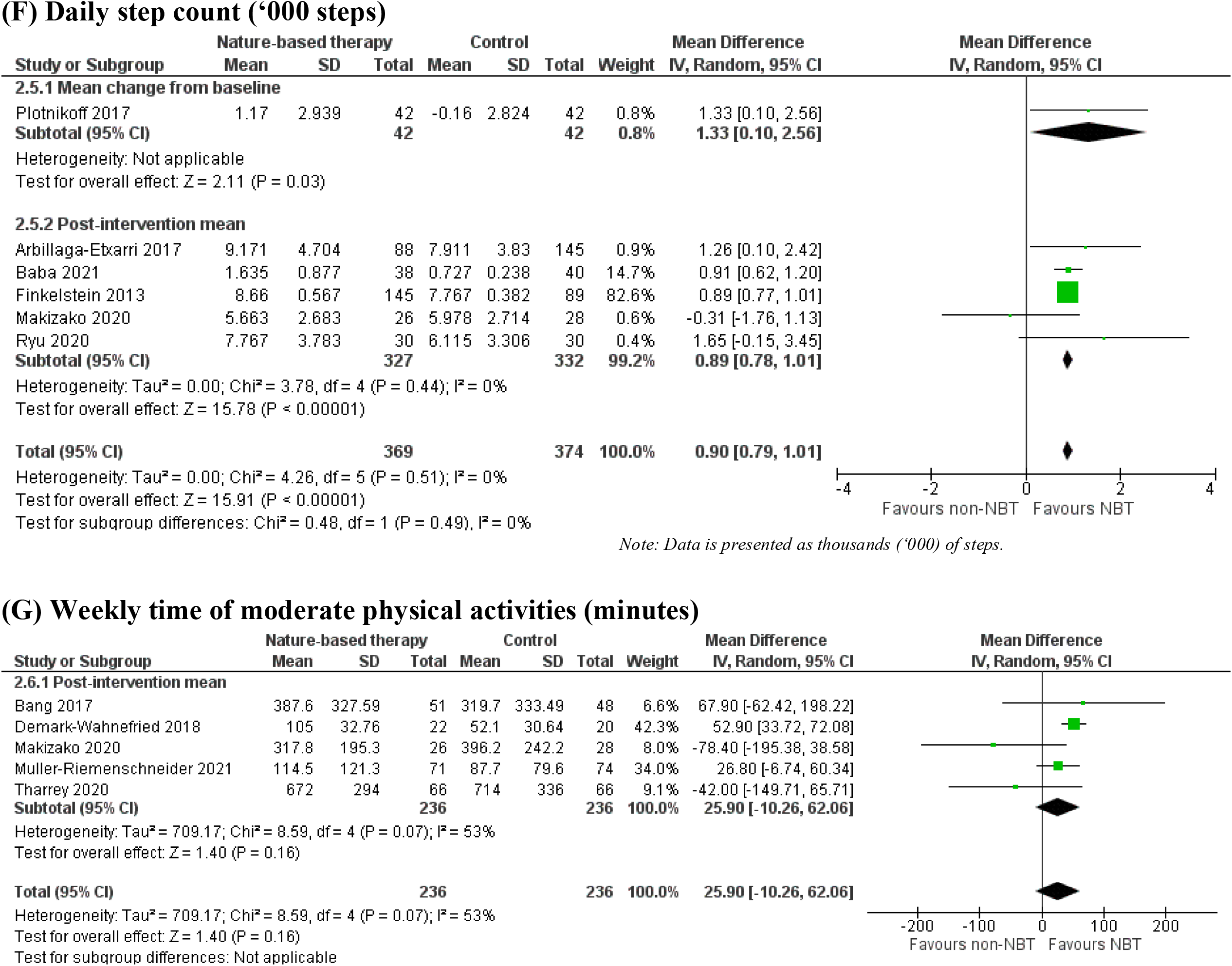
Forest plots of health outcomes.

Seven other studies, comprising five RCTs and two NRSs, were not included in the meta-analysis because of insufficient data or reasons related to study design (see Supplementary File S4). One study evaluated a clinic-community programme of organised games and sports at urban parks for obese children. The study reported a significant decrease in percentage of children classified as high or borderline blood pressure after 6 months, but no significant improvements in actual SBP or DBP percentile ^28^. Other studies reported that walking in forests or parks was linked to a higher decrease in SBP and DBP compared to control among elderly with hypertensions ^29–31^ and healthy adults ^31,32^. A cross-over trial reported improved blood pressure outcomes after walking in a green environment, but the improvement was not different from walking in a suburban environment ^33^.

#### 3.3.2. Depression and anxiety

Eleven RCTs and four NRSs contributed data to the meta-analysis (Figure 2C-D). The follow-up time ranged from 2 weeks to 1 year from baseline, except for one study ^34^ which followed up within 2 days from baseline. Compared to control, nature prescription programmes had a moderate-to-large effect on depression scores (SMD=0·5, 95% CI 0·2 to 0·8, I^2^=79%) and anxiety score (SMD=0·6, 95% CI 0·1 to 1·2, I^2^=90%). The most frequently-used tools were Beck Depression Inventory (n=5) for depression, and State-Trait Anxiety Inventory (n=4) or Hospital Anxiety and Depression Scale (n=4) for anxiety.

Five other studies, comprising four RCTs and one NRS, were not included in the meta-analysis because insufficient data was provided in the articles. All five studies evaluated horticulture therapies, and all reported that horticulture or gardening activities improved depression and anxiety symptoms among the elderly ^35^, stroke survivors ^36^ or military veterans ^37^ compared to baseline, but not significantly better than control. For psychiatric patients, Vujcic and colleagues ^38^ reported that horticulture therapy relieved stress but not depression nor anxiety.

#### 3.3.3. Physical activity levels

Seven RCTs and three NRSs contributed data to the meta-analysis (Figure 2E-F). The follow-up time ranged from 10 weeks to 1 year from baseline. Compared to control, nature prescription programmes resulted in a greater increase in daily step counts (MD = 900 steps, 95% CI 790-1010, I^2^=0%), but did not improve weekly time of moderate physical activities (MD = 25·9 minutes, 95% CI -10·3 to 62·1, I^2^=53%).

Six other studies, comprising three RCTs and three NRSs, were not included in the meta-analysis because of insufficient data or reasons related to study design (see Supplementary File S4). One study showed that officer workers taking lunch walks in a natural environment was more likely to achieve target step counts that those in a built environment ^39^. Similar benefits were observed in community gardening programme for obese adults ^40^ or farm-based day care for patients with dementia ^41^. In a study of cancer survivors, however, outdoor exercises did not have greater impact on long-term physical activities than indoor exercises ^42^. Among school students, nature-based activities did not increase moderate physical activity during play time more than playground sports ^43^. Razani and colleagues ^23^ reported that compared to park prescription alone, addition support in forms of text reminders and invitation to group nature outings resulted in a significant increase in park visits, but not levels of moderate physical activities.

## 4. DISCUSSION

The rising popularity of nature prescription programmes is a response to assumptions in healthcare challenged by COVID-19 and our ongoing climate crisis ^44,45^. Our scoping review identified a range of promising nature-based interventions that can be implemented as nature prescriptions. These interventions were demonstrated to be effective for various age groups, including children and the elderly, and targeted various health conditions, such as cardiovascular conditions, musculoskeletal disorders and psychiatric disorders. In addition, the nested meta-analyses on key outcomes demonstrated positive benefits on blood pressure, symptoms of depression and anxiety, and physical activity levels. This aligns with findings from studies on the effects of the nature environment on cardiometabolic health ^5^ and mental health ^14^.

The following key observations were made after examining the characteristics of these interventions, which can inform design of future nature prescription programmes. Firstly, these nature prescription programmes took place across diverse nature settings, including both green spaces and blue spaces. Green spaces can be urban landscape such as parks, botanic gardens, or nature environments tailored to the activities such as farms and gardens for horticulture, or forests for forest bathing. Secondly, nature prescription programmes can utilise a range of different activities suitable to the health conditions of the participants. Many of the included studies included multimodal interventions which incorporated a vigorous physical activity (e.g. walking, gardening) with a mindfulness-based activity (e.g. meditation, relaxation). Thirdly, beside health providers, social and community services were also effective channels to introduce participants to the intervention. Some studies were implemented as workplace programmes for knowledge-intensive or office workers. These institutions should be tapped on when designing future nature prescription programmes to maximise outreach and recruitment. Lastly, most of the included studies that reported positive impact on behavioural changes also demonstrated all three aspects of the SCT framework. This suggests the usefulness of the SC to guide the design of future prescription programmes.

Our review complements previous findings on nature prescriptions, which was limited to prescriptions dispensed in an outpatient setting ^13^. By using a broad scope, our review captured nature-based interventions that were dispensed outside the clinic setting and did not self-label as nature prescriptions, but nonetheless would be effective as one. Moreover, we conducted meta-analysis to quantify effectiveness of these interventions across physical, psychological and behavioural outcomes, demonstrating the holistic nature of nature prescription programmes in health promotion.

Our study was not without limitations. Since our primary aim is to conduct a scoping review on all potential nature-based interventions, our search strategy was designed to be generic. Therefore, we may miss some studies that feature unconventional nature-based therapies. In addition, as we only included studies reported in English, we may exclude relevant studies reported in other languages and introduce bias due to missing data, especially considering many studies are from East Asian countries (e.g. South Korea or Japan). Our data collection and risk-of-bias assessment was not conducted in duplicate, which potentially introduces some subjectivity.

Heterogeneity statistics from our meta-analysis suggests high degree of heterogeneity in true effects among our included studies, possibly due to different target populations, nature settings and activities featured in the intervention. Future studies are required to examine the varying effectiveness of nature-based prescriptions based on these factors. Moreover, a comparison of effectiveness on increasing physical activity levels based on different elements of the SCT will help identify factors that make a behavioural change programme successful. Most studies have moderate to high risk of bias, principally due to non-blinding nature of the interventions, small sample size and a lack of published documentations to rule out bias, such as an a priori analysis plan or protocol. This calls for future efforts to enhance the standards of reporting and conduct of trials in this area of research to improve the overall quality of evidence.

## 5. CONCLUSION

Nature prescription programmes are increasing in popularity around the world. A key impetus is for nature prescription programs to supplement health practitioner focus on biomedical options by attending to health and social needs that standard care cannot reach. Our review and meta-analysis concludes that present evidence indicates nature prescriptions can provide positive benefits on blood pressure, symptoms of depression and anxiety, and physical activity levels. Nature prescriptions should incorporate nature-based interventions, which can feature a range of natural settings and activities. Social and community channels should be utilised for outreach, in addition to health providers. The Social Cognitive Theory framework can be used to guide the design of an effective nature prescription programme.

## Supporting information

Supplementary File S1: Search strategy

Supplementary File S2: Risk of bias assessments

Supplementary File S3: Excluded studies with reasons

Supplementary File S4: Summary of intervention characteristics

Supplementary File S5: PRISMA checklist

Supplementary Figure S1: Distribution by year

Supplementary Figure S2: Distribution by location

## Data Availability

All data and materials used to generate the results are available in the manuscript and Supplementary Files, which can be accessed at doi.org/10.17605/OSF.IO/DKSJ9.

https://osf.io/dksj9/

## Contributor

P-Y.N: data curation (database search), formal analysis (study selection & data collection, ROB assessment, meta-analysis), writing - original draft

H.R-A: formal analysis (study selection & data collection)

X.F.: conceptualisation, funding acquisition, supervision, writing - review & editing

T.A-B: conceptualisation, funding acquisition, supervision, writing - review & editing

All authors had full access to all the data in the study and had final responsibility for the decision to submit for publication.

More than one author has directly accessed and verified the underlying data reported in the manuscript.

## Acknowledgement

This work was supported by the Hort Frontiers Green Cities Fund, part of the Hort Frontiers strategic partnership initiative developed by Hort Innovation, with co-investment from the University of Wollongong (UOW) Faculty of Social Sciences, the UOW Global Challenges initiative and contributions from the Australian Government (project number #GC15005). T.A-B. was supported by a National Health and Medical Research Council Boosting Dementia Research Leader Fellowship (#1140317). X.F. was supported by a National Health and Medical Research Council Career Development Fellowship (#1148792).

## Data availability statement

All data and materials used to generate the results are available in the manuscript and Supplementary Files.

