## Supplementary File S1: Search strategy for "Nature prescriptions: a scoping review with a nested meta-analysis"

**Ovid – Last date searched: July 28, 2021**

**1. Ovid MEDLINE(R) and Epub Ahead of Print, In-Process, In-Data-Review & Other Non-Indexed Citations, Daily and Versions(R) 1946 to July 27, 2021**

**2. Embase Classic+Embase 1947 to 2021 July 27**

**3. APA PsycInfo 1806 to July Week 3 2021**

| 1 | (naturerx or parksrx or parkrx or outdoorrx or nature-rx or park-rx or parks-rx or outdoor-rx).tw. |
| --- | --- |
| 2 | ((nature or park or social or green or garden* or outdoor) adj1 (prescription* or prescrib*)).tw. |
| 3 | ((outdoor play or outdoor activit* or (parks or park-based) or horticultur* or nature-based or nature-assisted or green space or greenspace or nature play or nature exposure or nature engagement or wander garden or forest bath* or forest walk* or shinrin-yoku) adj5 (health intervention* or clinical intervention* or medical intervention* or intervention* package* or referral)).ti,ab. |
| 4 | ((outdoor play or outdoor activit* or (parks or park-based) or horticultur* or nature-based or nature-assisted or green space or greenspace or nature play or nature exposure or nature engagement or wander garden or forest bath* or forest walk* or shinrin-yoku) adj5 (doctor* or physician* or clinician* or clinic or clinics or general practitioner* or health worker* or healthcare worker* or health care worker* or care provider* or medical provider* or health practitioner* or hcws)).ti,ab. |
| 5 | (nature therap* or nature-based therap* or nature-based intervention*).tw. |
| 6 | (park therap* or park-based therap*).tw. |
| 7 | (green therap* or green care).tw. |
| 8 | (farm care or care farm* or farm therapy or therapeutic farm*).tw. |
| 9 | (therapeutic horticulture or therapeutic gardening).tw. |
| 10 | ((green or blue) adj1 (gym or gyms or exercis*)).tw. |
| 11 | ((environment* or outdoor or nature* or biodiversity or conservation) adj1 (volunteer or volunteering)).ti,ab. |
| 12 | ((environment* or outdoor or nature*) adj2 (exercis* or walks or walking or walk or guided) adj2 (group or groups or program* or guided)).tw. |
| 13 | (nature play or wild play).tw. |
| 14 | ((nature or outdoor) adj2 (arts or art or crafts or craft or painting or drawing or photography or poetry) adj5 (therap* or health or intervention*)).ti,ab. |
| 15 | (forest bath* or forest therapy or shinrin-yoku or (forest* adj3 (walk or walks or walking or spend* time or meditat* or relax*))).ti,ab. |
| 16 | ((outdoor play or outdoor activit* or (parks or park-based) or horticultur* or nature-based or nature-assisted or green space or greenspace or nature play or natural exposure or nature engagement or wander garden or forest bath* or forest walk* or shinrin-yoku) adj5 therap*).ti,ab. |
| 17 | ((outdoor play or outdoor activit* or (parks or park-based) or horticultur* or nature-based or nature-assisted or green space or greenspace or nature play or nature exposure or nature engagement or wander garden or forest bath* or forest walk* or shinrin-yoku) adj5 treatment*).ti,ab. |
| 18 | (wilderness adj2 (program* or therapy)).tw. |
| 19 | residential retreat*.ti,ab. |
| 20 | (Ecotherapy or eco-therapy).tw. |
| 21 | (Eco-education or ecoeducation).tw. |
| 22 | (outdoor education or outdoor classroom* or outdoor curriculum or outdoor school* or outdoor learning or forest school*).ti,ab. |
| 23 | (eco-psychology or ecopsychology).tw. |
| 24 | ((green space or greenspace or outdoor or (nature adj3 (contact or expos* or play or visit* or activit* or time spent)) or forest* or parks or park or garden* or open space* or public space* or garden*) and (nudge or nudges or nudging)).ti,ab. |
| 25 | ((green space or greenspace or outdoor or (nature adj3 (contact or expos* or play or visit* or activit* or time spent)) or forest* or parks or park or garden* or open space* or public space* or garden*) and (promot* or incentivi* or motivat*)).ti. |
| 26 | ((green space or greenspace or outdoor or (nature adj3 (contact or expos* or play or visit* or activit* or time spent)) or forest* or parks or park or garden* or open space* or public space* or garden*) adj5 ((increas* or promot* or motivat* or incentiv*) adj2 (visit* or play time or activit* or time spent))).ti. |
| 27 | ((green space or greenspace or outdoor or (nature adj3 (contact or expos* or play or visit* or activit* or time spent)) or forest* or parks or park or garden* or open space* or public space* or garden*) and ((program* or intervention* package* or strateg*) adj5 (visit* or play time or activit* or time spent))).ti,ab. |
| 28 | ((green space or greenspace or outdoor or (nature adj3 (contact or expos* or play or visit* or activit* or time spent)) or forest* or parks or park or garden* or open space* or public space* or garden*) and ((coach* or trainer* or personalised or personalized) adj5 (visit* or play time or activit* or time spent))).ti,ab. |
| 29 | ((green space or greenspace or outdoor or (nature adj3 (contact or expos* or play or visit* or activit* or time spent)) or forest* or parks or park or garden* or open space* or public space* or garden*) and ((target* or progress or goal) adj2 (tracking or self-monitor* or monitor* or setting) adj5 (visit* or play time or activit* or time spent))).ti,ab. |
| 30 | ((green space or greenspace or outdoor or (nature adj3 (contact or expos* or play or visit* or activit* or time spent)) or forest* or parks or park or garden* or open space* or public space* or garden*) and ((feedback* or prompts or cues) adj5 (visit* or play time or activit* or time spent))).ti,ab. |
| 31 | ((green space or greenspace or outdoor or (nature adj3 (contact or expos* or play or visit* or activit* or time spent)) or forest* or parks or park or garden* or open space* or public space* or garden*) and (social support adj5 (visit* or play time or activit* or time spent))).ti,ab. |
| 32 | ((green space or greenspace or outdoor or (nature adj3 (contact or expos* or play or visit* or activit* or time spent)) or forest* or parks or park or garden* or open space* or public space* or garden*) and (counsel* adj5 (visit* or play time or activit* or time spent))).ti,ab. |
| 33 | ((green space or greenspace or outdoor or (nature adj3 (contact or expos* or play or visit* or activit* or time spent)) or forest* or parks or park or garden* or open space* or public space* or garden*) and ((pamphlet* or brochure* or website* or hotline* or education* platform*) adj5 (visit* or play time or activit* or time spent))).ti,ab. |
| 34 | ((green space or greenspace or outdoor or (nature adj3 (contact or expos* or play or visit* or activit* or time spent)) or forest* or parks or park or garden* or open space* or public space* or garden*) and ((mobile or phone* or smartphone* or digital or telehealth or messag*) adj5 (visit* or play time or activit* or time spent))).ti,ab. |
| 35 | ((green space or greenspace or outdoor or (nature adj3 (contact or expos* or play or visit* or activit* or time spent)) or forest* or parks or park or garden* or open space* or public space* or garden*) and ((apps or app or app-based or pedometer) adj5 (visit* or play time or activit* or time spent))).ti,ab. |
| 36 | ((green space or greenspace or outdoor or (nature adj3 (contact or expos* or play or visit* or activit* or time spent)) or forest* or parks or park or garden* or open space* or public space* or garden*) and ((transport* or meal* or lunch or dinner) adj5 (visit* or play time or activit* or time spent))).ti,ab. |
| 37 | ((green space or greenspace or outdoor or (nature adj3 (contact or expos* or play or visit* or activit* or time spent)) or forest* or parks or park or garden* or open space* or public space* or garden*) and ((fees or cost* or waive* or discount* or price* or subsidi* or payment* or financial or monetary or voucher* or reimburse*) adj5 (visit* or play time or activit* or time spent))).ti,ab. |
| 38 | ((green space or greenspace or outdoor or (nature adj3 (contact or expos* or play or visit* or activit* or time spent)) or forest* or parks or park or garden* or open space* or public space* or garden*) and ((group or groups or group-based) adj5 (visit* or play time or activit* or time spent))).ti,ab. |
| 39 | ((green space or greenspace or outdoor or (nature adj3 (contact or expos* or play or visit* or activit* or time spent)) or forest* or parks or park or garden* or open space* or public space* or garden*) and (community* adj5 (visit* or play time or activit* or time spent))).ti,ab. |
| 40 | or/1-39 |
| 41 | 40 and randomized controlled trial.pt. |
| 42 | 40 and controlled clinical trial.pt. |
| 43 | 40 and (randomized or randomised or randomly).ab. |
| 44 | 40 and (placebo or control group or experiment group or intervention group).ab. |
| 45 | 40 and clinical trials as topic.sh. |
| 46 | 40 and (experiment or trial).ti. |
| 47 | 40 and (natural experiment* or quasi-experiment* or experimental).ab. |
| 48 | 40 and (crossover trial or (controlled adj4 trial) or (random* adj4 trial)).ab. |
| 49 | or/41-48 |
| 50 | 49 not (exp animals/ not humans.sh.) |
| 51 | 50 not (wastewater or waste-water or mosquito* or water quality or water pollution or pesticide* or outdoor clinic*).tw. |
