## Supplementary File S3: Excluded studies with reasons for "Nature prescriptions: a scoping review with a nested meta-analysis"

**Supplementary File S3. List of excluded full-texts**

| **Author/Year** | **Full title** | **Reason for exclusion** |
| --- | --- | --- |
| Duvall_2012 | A comparison of engagement strategies for encouraging outdoor walking | Not a nature intervention |
| ISRCTN04540006_2008 | A feasibility study to analyse the psychological benefits of green exercise (GE) in comparison to cognitive behavioural therapy (CBT) with patients with mild to moderate depression | Protocol without published results |
| Li_2011 | A multilevel intervention for HIV-affected families in China: together for Empowerment Activities (TEA) | Not a nature intervention |
| ACTRN12618002019280_2018 | A physical activity program for women living with metastatic breast cancer | Not a nature intervention |
| Lee_2018 | A pilot study: horticulture-related activities significantly reduce stress levels and salivary cortisol concentration of maladjusted elementary school children | Not a nature intervention |
| Wolfenden_2016 | A randomised controlled trial of multiple periods of outdoor free-play to increase moderate-to-vigorous physical activity among 3 to 6 year old children attending childcare: study protocol | School programmes with outdoor play, no clear nature focus |
| ISRCTN51290984_2018 | A research study in Bosnia and Herzegovina to test an intervention called Volunteer Support, designed to improve care for people living in the community with severe mental illness | Not a nature intervention |
| Logan_2003 | A study of interventions and related outcomes in a randomized controlled trial of occupational therapy and leisure therapy for community stroke patients | Not a nature intervention |
| JPRN-UMIN000035037_2018 | A study on psychological and fatigue recovery effect of horticultural therapy using medicinal crops and rural area walking | Protocol without published results |
| Brussoni_2021 | A web-based and in-person risk reframing intervention to influence mothers' tolerance for, and parenting practices associated with, children's outdoor risky play: randomized controlled trial | Not a nature intervention |
| Drum_2016 | Acute effects of walking at moderate normobaric hypoxia on gait and balance performance in healthy community-dwelling seniors: a randomized controlled crossover study | Not a nature intervention |
| Liu_2020 | Air Pollution, Physical Activity, and Cardiovascular Function of Patients With Implanted Cardioverter Defibrillators: a Randomized Controlled Trial of Indoor Versus Outdoor Activity | Not a nature intervention |
| Johnstone_2019 | An active play intervention to improve physical activity and fundamental movement skills in children of low socioeconomic status: feasibility cluster randomised controlled trial | School programmes with outdoor play, no clear nature focus |
| ACTRN12614000850673_2014 | An educational intervention to promote healthy lifestyles in preschool aged children | Protocol without published results |
| NCT04543058_2020 | BeatPark: self-rehabilitation in Walking With Parkinson's Disease | Ongoing study |
| NCT02824861_2016 | Bemobile Intervention to Support Physical Activity in Cancer Survivors | Protocol without published results |
| Merom_2016 | Cognitive benefits of social dancing and walking in old age: the dancing mind randomized controlled trial | Not a nature intervention |
| NCT04725487_2021 | Community With Immigrants - a Step on the Road to Employment | Ongoing study |
| ISRCTN11607781_2017 | Does participating in organised active play sessions increase children's physical activity levels? | School programmes with outdoor play, no clear nature focus |
| NCT04624932_2020 | Early Childhood Outside (ECO) - Randomized Controlled Trial Study | Ongoing study |
| RBR-9zgxzd_2018 | Effect of different types of physical exercise in SUS patients with depression | Protocol without published results |
| NCT04587765_2020 | Effect of Extra-curricular Sports Activities After School on Primary School Children's Academic Performance | School programmes with outdoor play, no clear nature focus |
| ChiCTR-TRC-14004735_2014 | Effect of forest bathing on patients with stable chronic obstructive pulmonary disease | Protocol without published results |
| Iwamoto_1998 | Effect of increased physical activity on bone mineral density in postmenopausal osteoporotic women | Protocol without published results |
| ISRCTN12698269_2016 | Effectiveness of the run-a-mile intervention | School programmes with outdoor play, no clear nature focus |
| VanDyck_2016 | Effectiveness of the self-regulation eHealth intervention 'MyPlan1.0.' on physical activity levels of recently retired Belgian adults: a randomized controlled trial | Not a nature intervention |
| NCT02429544_2015 | Effects of a Group Residential Retreat on Cancer Outcomes | Protocol without published results |
| JPRN-UMIN000028265_2017 | Effects of Adachi Rehabilitation Programme (ARP) for older adults with long term care level 1 to 3: a randomised controlled trial | Duplicate / already included |
| JPRN-UMIN000033152_2018 | Effects of agricultural and horticultural activity | Protocol without published results |
| ChiCTR2000034495_2020 | Effects of an outdoor multisurface terrain fall prevention exercise program on the walking and balance abilities in the older adults: a randomized controlled trial | Not a nature intervention |
| NCT04936672_2021 | Effects of Brisk Walking Combined With Tai Chi Chuan on Health-Related Physical Fitness and Selected Health Parameters Among Older Chinese Women | Ongoing study |
| NCT03045718_2017 | Effects of Horticultural Therapy on Elderly at Risk of Cognitive Decline | Protocol without published results |
| RBR-7b8f2c_2018 | Effects of physical exercise programs performed at outdoor fitness gym in hypertensive individuals | Protocol without published results |
| NCT04780646_2021 | Effects of Urban Nature on Stress and Quality of Life | Ongoing study |
| DRKS00024152_2021 | Efficacy and Cost Effectiveness of the Implementation of a Supervised, Transdiagnostic Group-based Exercise Intervention Named ImPuls for Patients With Major Depression, Panic Disorder, Agoraphobia, Post-traumatic Stress Disorder and Nonorganic Insomnia | Ongoing study |
| Chu_2019 | Efficacy of a horticultural activity program for reducing depression and loneliness in older residents of nursing homes in Taiwan | Not a nature intervention |
| deVries_2015 | Efficacy of an exercise intervention for employees with work-related fatigue: study protocol of a two-arm randomized controlled trial | Not a nature intervention |
| NCT04712968_2021 | Efficacy of Daylight as Adjunctive Treatment in Patients With Depression | Not a nature intervention |
| ISRCTN58237124_2015 | Exercise intensity and diabetes type 2 | Protocol without published results |
| NCT02371954_2015 | Exercise to Prevent Depression and Anxiety in Older Hispanics | Protocol without published results |
| Arredondo_2015 | Fe en Accion/Faith in Action: design and implementation of a church-based randomized trial to promote physical activity and cancer screening among churchgoing Latinas | Protocol without published results |
| McGarty_2021 | Feasibility of the Go2Play Active Play intervention for increasing physical and social development in children with intellectual disabilities | School programmes with outdoor play, no clear nature focus |
| Warber_2011 | Healing the heart: a randomized pilot study of a spiritual retreat for depression in acute coronary syndrome patients | Inappropriate comparison |
| NCT00994084_2009 | Healthy Kids-Houston: a Community Childhood Obesity Intervention Program | School programmes with outdoor play, no clear nature focus |
| Verra_2011 | Horticultural therapy for patients with chronic musculoskeletal pain: results of a pilot study | Wrong publication type |
| ACTRN12619001320145_2019 | How does a Movement Oriented Games Based Assessment (MOGBA) programme impact upon the movement competence, physical activity levels and self-perceptions of children aged 8-12 years of age? | Not a nature intervention |
| JPRN-UMIN000014992_2014 | Impact of a life style intervention in incident and prevalence of overweight and obesity among secondary school children in Hanoi | School programmes with outdoor play, no clear nature focus |
| Rice_1998 | Impact of horticultural therapy on psychosocial functioning among urban jail inmates | Wrong publication type |
| Reddy_2021 | Impact of neighborhood factors on physical activity among urban African American women with asthma | Not a behavioural change intervention |
| NCT04593576_2020 | Impact of Sports Activity on Pragmatic Skills of Children | Protocol without published results |
| NCT03568682_2018 | Improving Antiretroviral Therapy Adherence Through Urban Gardening and Nutritional Counseling in the Dominican Republic | Protocol without published results |
| Anandh_2020 | Influence of dual task training in indoor versus outdooenvironment on physical function and social activity among elderly | Not a nature intervention |
| Zhang_2021 | Intervention Effect of Research-based Psychological Counseling on Adolescents' Mental Health during the COVID-19 Epidemic | Not a nature intervention |
| Wadsworth_2020 | Intervention Strategies to Elicit MVPA in Preschoolers during Outdoor Play | School programmes with outdoor play, no clear nature focus |
| JPRN-UMIN000008936_2012 | Intervention study of psychotherapy on elderly women who live in disaster area | Protocol without published results |
| Rodriguez-Romero_2020 | Intervention to reduce perceived loneliness in community-dwelling older people | Not a nature intervention |
| ACTRN12617000311358_2017 | KOALA Healthy Life Program for promoting healthy lifestyles and weight in children with obesity | Protocol without published results |
| NCT03673722_2018 | Mediterranean Diet, Exercise and Dementia Risk in UK Adults | Not a nature intervention |
| ACTRN12619000936123_2019 | Motivational interviewing to increase walking in community-dwelling older adults after hip fracture | Ongoing study |
| JPRN-UMIN000013342_2014 | Multicenter clinical study on Blue Exercise for type2 diabetes patients | Protocol without published results |
| NCT04642235_2020 | Nature and Well-Being Project | Ongoing study |
| NCT03501953_2018 | Neurophysiological Mechanisms of Physical Activity Interventions in Unique Environments | Protocol without published results |
| Westmoreland_1975 | Outdoor activity group experience and group counseling with institutionalized children and adolescents | Complex outdoor programme |
| Levinger_2018 | Outdoor physical activity for older people-the senior exercise park: current research, challenges and future directions | Not an interventional study |
| DRKS00010977_2016 | Outdoor walking training versus cycle ergometer training during inpatient rehabilitation in COPD patients GOLD stages III to IV. A feasibility randomized controlled trial | Not a nature intervention |
| Eskicioglu_2014 | Peer mentoring for type 2 diabetes prevention in First Nations children | Not a nature intervention |
| King_2017 | Preserving older adults' routine outdoor activities in contrasting neighborhood environments through a physical activity intervention | Not a nature intervention |
| DRKS00015188_2018 | Prevention by lay-assisted Outdoor-Walking in the Elderly at Risk (The POWER Study): a randomized controlled trial | Ongoing study |
| DRKS00000745_2011 | Prevention of falls and fall-related fractures in elderly > 65 at risk of falls by the means of regular physical activity - a pilot study | Protocol without published results |
| NCT00775216_2008 | Project Diabetes: prescription for Healthy Living | Protocol without published results |
| NCT02715544_2016 | Promoting Healthy Eating and Active Playtime by Connecting Preschool Children to Nature | Protocol without published results |
| ISRCTN37185489_2010 | Recovery through healthy living in acute psychiatric setting | Protocol without published results |
| IRCT2014092113850N2_2015 | Recreation Therapy on Depression in Elderly | Protocol without published results |
| Logan_2014 | Rehabilitation aimed at improving outdoor mobility for people after stroke: a multicentre randomised controlled study (the Getting out of the House Study) | Not a nature intervention |
| Wallner_2018 | Reloading pupils' batteries: impact of green spaces on cognition and wellbeing | Inappropriate comparison |
| VanDyck_2019 | Results of MyPlan 2.0 on Physical Activity in Older Belgian Adults: randomized Controlled Trial | Not a nature intervention |
| ACTRN12621000317897_2021 | Running for health: a pilot randomized controlled trial investigating the effect of RaceRunning training on cardiovascular health in children and youth with cerebral palsy | Protocol without published results |
| Ceci_1991 | Self-monitored exercise at three different RPE intensities in treadmill vs field running | Not a nature intervention |
| He_2019 | Shanghai Time Outside to Reduce Myopia trial: design and baseline data | Ongoing study |
| JPRN-UMIN000005004_2011 | Spiritual care of cancer patients by integrated medicine in a forest park | Single-group design |
| NCT04817995_2021 | Stress Recovery Program Forest for Healthcare Staff: a Randomized Controlled Trial | Protocol without published results |
| NCT01762553_2013 | TEA for Families and Children: a Randomized Intervention Trial | Not a nature intervention |
| Wolf_1993 | The Atlanta FICSIT study: two exercise interventions to reduce frailty in elders | Not a nature intervention |
| ChiCTR2100042770_2021 | The construction and feasibility evaluation of horticultural therapy intervention scheme for elderly breast cancer patients based on happiness PERMA theory | Protocol without published results |
| NCT04846790_2021 | The Effect of a Combined Nature-based and Virtual Mindfulness Intervention on Perceived Stress in Healthcare Workers | Ongoing study |
| Volders_2020 | The Effect of Active Plus, a Computer-Tailored Physical Activity Intervention, on the Physical Activity of Older Adults with Chronic Illness(es)-A Cluster Randomized Controlled Trial | Not a nature intervention |
| Pirzadeh_2017 | The effect of exercise on menopausal symptoms in postmenopausal women | Wrong publication type |
| Luk_2011 | The effect of horticultural activities on agitation in nursing home residents with dementia | Wrong publication type |
| Zachor_2016 | The effectiveness of an outdoor adventure programme for young children with autism spectrum disorder: a controlled study | Duplicate / already included |
| NCT01799681_2013 | The Effects of the Hopeful Outdoor Parkinson Exercise (HOPE) Program on Improving Balance Performance in Parkinsonian Non-fallers and Single Fallers | Duplicate / already included |
| Driediger_2019 | The Impact of Shorter, More Frequent Outdoor Play Periods on Preschoolers' Physical Activity during Childcare: a Cluster Randomized Controlled Trial | School programmes with outdoor play, no clear nature focus |
| Tillekeratne_2017 | The importance of coaching and mentoring, post nutrition education for sustained behaviour changes-the USAID solid approach | No appropriate outcomes measured |
| NCT01574352_2012 | The Odense Overweight Intervention Study | Complex outdoor programme |
| NCT01575262_2012 | The Physical Activity Loyalty Card Scheme | Protocol without published results |
| NCT02980445_2016 | Time Outdoors as an Intervention for Myopia in Children | Protocol without published results |
| Tandon_2019 | Two Approaches to Increase Physical Activity for Preschool Children in Child Care Centers: a Matched-Pair Cluster-Randomized Trial | School programmes with outdoor play, no clear nature focus |
| Arbillaga-Etxarri_2016 | Validation of walking trails for the urban training of chronic obstructive pulmonary disease patients | Not a behavioural change intervention |
| NCT03803085_2019 | Walking Competition to Enhance Daily Physical Activity and Social Engagement Among Older Adults | Not a nature intervention |
| ISRCTN34755197_2012 | Watch Me Grow: a garden-based pilot intervention to increase fruit and vegetable intake in preschoolers | No appropriate outcomes measured |
| Barber_2013 | "Pre-schoolers in the playground" an outdoor physical activity intervention for children aged 18 months to 4 years old: study protocol for a pilot cluster randomised controlled trial. | Protocol without published results |
| Miyazaki_2011 | [Preventive medical effects of nature therapy] | Not in English |
| Tsunetsugu_2011 | [Psychological relaxation effect of forest therapy: results of field experiments in 19 forests in Japan involving 228 participants] | Not in English |
| Morita_2011 | A before and after comparison of the effects of forest walking on the sleep of a community-based sample of people with sleep complaints. | Single-group design |
| McCluskey_2016 | A behavior change program to increase outings delivered during therapy to stroke survivors by community rehabilitation teams: the Out-and-About trial | Not a nature intervention |
| Temple_2014 | A Built Environmental Intervention and a Combination Built Environmental and Cognitive/Behavioral Intervention to Increase Individual Physical Activity in 3 to 5- Year-Old Children. | Wrong publication type |
| Booth_2020 | A citizen science study of short physical activity breaks at school: Improvements in cognition and wellbeing with self-paced activity | School programmes with outdoor play, no clear nature focus |
| Knox_2009 | A cross-curricular physical activity intervention to combat cardiovascular disease risk factors in 11-14 year olds: 'activity knowledge circuit'. | School programmes with outdoor play, no clear nature focus |
| Morimoto_2008 | A forest bathing trip increases human natural killer activity and expression of anti-cancer proteins in female subjects | Single-group design |
| NCT00974727_2009 | A Gardening Program to Assess Unhealthy Lifestyle Contributions to Summer Weight Gain in Children | Protocol without published results |
| Pfeiffer_2001 | A green prescription study: does written exercise prescribed by a physician result in increased physical activity among older adults? | Not a nature intervention |
| Lim_2020 | A guide to nature immersion: Psychological and physiological benefits | Inappropriate comparison |
| Chiumento_2018 | A haven of green space: learning from a pilot pre-post evaluation of a school-based social and therapeutic horticulture intervention with children | Single-group design |
| Chiumento_2018 | A haven of green space: learning from a pilot pre-post evaluation of a school-based social and therapeutic horticulture intervention with children. | Duplicate / already included |
| Houser_2019 | A Loose Parts Randomized Controlled Trial to Promote Active Outdoor Play in Preschool-aged Children: Physical Literacy in the Early Years (PLEY) Project. | School programmes with outdoor play, no clear nature focus |
| Logan_2012 | A multi-centre randomised controlled trial of rehabilitation aimed at improving outdoor mobility for people after stroke: study protocol for a randomised controlled trial | Not a nature intervention |
| Saez_2021 | A multicomponent program to improve self-concept and self-esteem among intimate partner violence victims: a study protocol for a randomized controlled pilot trial | Ongoing study |
| vanderKolk_2014 | A personalized coaching program increases outdoor activities and physical fitness in sedentary Parkinson patients; a post-hoc analysis of the ParkFit trial | Not a nature intervention |
| ACTRN12620000266965_2020 | A pilot gender-sensitised lifestyle intervention for overweight men targeted at physical activity, diet, and mental health | Protocol without published results |
| Stowell_2018 | A pilot horticultural therapy program serving veterans with mental health issues: Feasibility and outcomes | Single-group design |
| Patten_2017 | A pilot study of children's physical activity levels during imagination-based mobile games | Not a nature intervention |
| Alhassan_2013 | A Pilot Study to Examine the Effect of Additional Structured Outdoor Playtime on Preschoolers' Physical Activity Levels | School programmes with outdoor play, no clear nature focus |
| Gonzalez_2011 | A prospective study of existential issues in therapeutic horticulture for clinical depression. | Single-group design |
| Goyder_2014 | A randomised controlled trial and cost-effectiveness evaluation of 'booster' interventions to sustain increases in physical activity in middle-aged adults in deprived urban neighbourhoods. | Protocol without published results |
| ACTRN12612000937819_2012 | A Randomised Controlled Trial of Co-ordinated Lifestyle Advice in Patients Admitted with an Acute Coronary Syndrome | Protocol without published results |
| Palsdottir_2017 | A randomized controlled study on nature-based rehabilitation for poststroke fatigue-Â« the nature stroke studyÂ» (NASTRU) | Duplicate / already included |
| Sobko_2016 | A randomized controlled trial for families with preschool children - promoting healthy eating and active playtime by connecting to nature. | No appropriate outcomes measured |
| Reed_2013 | A repeated measures experiment of green exercise to improve self-esteem in UK school children | No health outcomes measured |
| Fung_2015 | A study of the effectiveness of a horticultural therapy group on the emotional health of cancer patients | Wrong publication type |
| Cooke_2020 | A trajectory analysis of daily step counts during a physician-delivered intervention | Not a nature intervention |
| NCT03339440_2017 | A Trial of an Integrated Clinic-community Intervention in Children and Adolescents With Obesity (Hearts and Parks) | Not a nature intervention |
| Grede_2021 | A volunteer-supported walking programme to improve physical function in older people (the POWER Study): study protocol for a randomised controlled trial | Ongoing study |
| DRKS00007549_2015 | A year in the forest: the influence of nature-orientated and high activation school teaching on biological indicators of stress resilience | Not an interventional study |
| NCT03786198_2018 | Activity Program During Aromatase Inhibitor Therapy | Not a nature intervention |
| Li_2011 | Acute effects of walking in forest environments on cardiovascular and metabolic parameters | Single-group design |
| Lacharite-Lemieux_2014 | Adherence to exercise and metabolic improvements in postmenopausal women: Comparison between outdoor and indoor training | Duplicate / already included |
| Rogerson_2020 | Affective Outcomes of Group versus Lone Green Exercise Participation | Inappropriate comparison |
| Niedermeier_2017 | Affective responses in mountain hiking-A randomized crossover trial focusing on differences between indoor and outdoor activity | Complex outdoor programme |
| Luckner_1989 | Altering locus of control of individuals with hearing impairments by outdoor-adventure courses. | Complex outdoor programme |
| Horiuchi_2015 | An effective strategy to reduce blood pressure after forest walking in middle-aged and aged people | Single-group design |
| Bayer_2019 | An eight-week forest yoga intervention for chronic pain: Effect on pain interference, pain severity, and psychological outcomes. | Wrong publication type |
| Pringle_2009 | An evaluation of the Local Exercise Action Pilots and impact on moderate physical activity | Not a nature intervention |
| Podina_2017 | An evidence-based gamified mHealth intervention for overweight young adults with maladaptive eating habits: study protocol for a randomized controlled trial | Not a nature intervention |
| Kang_2010 | An integrated dementia intervention for Korean older adults | Not a nature intervention |
| NCT01373307_2011 | An Intergenerational Community Based Participatory Research (CBPR) Intervention to Reduce Appalachian Health Disparities | Protocol without published results |
| Rosenberg_2014 | An Outdoor Adventure Program for Young Adults with Cancer: Positive Effects on Body Image and Psychosocial Functioning. | Complex outdoor programme |
| Thomson_2020 | Art, nature and mental health: assessing the biopsychosocial effects of a 'creative green prescription' museum programme involving horticulture, artmaking and collections | Single-group design |
| Bunpo_2016 | Ascorbic acid supplementation does not alter oxidative stress markers in healthy volunteers engaged in a supervised exercise program | Not a nature intervention |
| Torres_2017 | Assessing the effect of physical activity classes in public spaces on leisure-time physical activity: "Al Ritmo de las Comunidades" A natural experiment in Bogota, Colombia | Duplicate / already included |
| Torres_2017 | Assessing the effect of physical activity classes in public spaces on leisure-time physical activity: "Al Ritmo de las Comunidades" A natural experiment in Bogota, Colombia. | Inappropriate comparison |
| Yang_2014 | Beneficial effect of forest bathing for elderly patients with stable chronic obstructive pulmonary disease | Duplicate / already included |
| Jia_2016 | Beneficial effect of forest bathing on elderly patients with chronic obstructive pulmonary disease | Duplicate / already included |
| Gagliardi_2020 | Benefits for older people engaged in environmental volunteering and socializing activities in city parks: Preliminary results of a program in Italy | Single-group design |
| Zeng_2020 | Benefits of a Three-Day Bamboo Forest Therapy Session on the Physiological Responses of University Students | No appropriate outcomes measured |
| Freeman_2017 | Benefits of walking and solo experiences in UK wild places. | Inappropriate comparison |
| Sturm_2020 | Big Smile, Small Self: awe Walks Promote Prosocial Positive Emotions in Older Adults | Inappropriate comparison |
| Mawani_2021 | Building Roads Together: a peer-led, community-based walking and rolling peer support program for inclusion and mental health | Qualitative study |
| Merom_2009 | Can a motivational intervention overcome an unsupportive environment for walking--findings from the Step-by-Step Study | Not a nature intervention |
| Walasek_2021 | Can a 'rewards-for-exercise app' increase physical activity, subjective well-being and sleep quality? An open-label single-arm trial among university staff with low to moderate physical activity levels | Not a nature intervention |
| ISRCTN68006446_2019 | Can adventure learning improve studentsa[Euro sign][TM] skills, behaviour and academic results? | Protocol without published results |
| Jiang_2012 | Can exercise enhance smoking cessation outcomes? A pragmatic randomized controlled trial (fit2quit Study) | Not a nature intervention |
| Pasanen_2018 | Can nature walks with psychological tasks improve mood, self-reported restoration, and sustained attention? Results from two experimental field studies. | Inappropriate comparison |
| NCT04864574_2021 | Childcare Outdoor Learning Environments as Active Food Systems | School programmes with outdoor play, no clear nature focus |
| Razani_2019 | Clinic and park partnerships for childhood resilience: A prospective study of park prescriptions. | Single-group design |
| Bammann_2018 | Cluster-randomised trial on participatory community-based outdoor physical activity promotion programs in adults aged 65-75 years in Germany: protocol of the OUTDOOR ACTIVE intervention trial | Protocol without published results |
| Bammann_2018 | Cluster-randomised trial on participatory community-based outdoor physical activity promotion programs in adults aged 65-75 years in Germany: Protocol of the OUTDOOR ACTIVE intervention trial 11 Medical and Health Sciences 1117 Public Health and Health Se | Protocol without published results |
| Elliot_2018 | Combined diet and physical activity is better than diet or physical activity alone at improving health outcomes for patients in New Zealand's primary care intervention | Not a nature intervention |
| Liden_2016 | Combining garden therapy and supported employment - a method for preparing women on long-term sick leave for working life | Inappropriate comparison |
| NCT03089177_2017 | Community Activation for Prevention (CAPs): a Study of Community Gardening | Ongoing study |
| Dillon_2020 | Community gardening: Stress, well-being, and resilience potentials | Not an interventional study |
| McCluskey_2010 | Community occupational therapists and physiotherapists delivered 15% more evidence-based outdoor journey sessions to peoplewith stroke after audit and feedback about their practice | Wrong target population |
| Jarrott_2010 | Comparing responses to horticultural-based and traditional activities in dementia care programs | Not a nature intervention |
| IRCT20151118025105N3_2019 | Comparing the effects of Exergaming with brisk walking in older adults | Not a nature intervention |
| Tharrey_2021 | Correction to: Improving lifestyles sustainability through community gardening: results and lessons learnt from the JArDinS quasi-experimental study. | Duplicate / already included |
| Lee_2019 | Design and methodology of a cluster-randomized trial in early care and education centers to meet physical activity guidelines: sustainability via Active Garden Education (SAGE) | School programmes with outdoor play, no clear nature focus |
| O'Brien_2017 | Developing a growth mindset through outdoor personal development: Can an intervention underpinned by psychology increase the impact of an outdoor learning course for young people? | Complex outdoor programme |
| deHeer_2019 | Development of a culturally relevant physical activity intervention for Navajo cancer survivors. | Ongoing study |
| NCT02215239_2014 | Development of Workplace Physical Activity Promotion Models in Taiwan | Protocol without published results |
| Sobko_2020 | Does connectedness to nature improve the eating behaviours of pre-schoolers? Emerging evidence from the Play&Grow randomised controlled trial in Hong Kong | No appropriate outcomes measured |
| Xie_2021 | Dose-response effect of a large-scale greenway intervention on physical activities: The first natural experimental study in China. | Not a behavioural change intervention |
| Derose_2019 | Eat, Pray, Move: a Pilot Cluster Randomized Controlled Trial of a Multilevel Church-Based Intervention to Address Obesity Among African Americans and Latinos | No appropriate outcomes measured |
| RBR-5db955_2020 | Effect of exercise training on clinical markers ,quality of life and mental health in women with polycystic ovary syndrome | Ongoing study |
| IRCT20180505039538N1_2018 | Effect of group walking program on social physical anxiety and eating disorders | Protocol without published results |
| Navalta_2021 | Effect of exercise in a desert environment on physiological and subjective measures | Inappropriate comparison |
| NCT04592770_2020 | Effect of Exercise on Post-traumatic Growth Among Health Care Providers With Post-traumatic Stress | Ongoing study |
| Lee_2011 | Effect of forest bathing on physiological and psychological responses in young Japanese male subjects | Single-group design |
| NCT02225847_2014 | Effect of Gardening on Brain Activity | Protocol without published results |
| Yang_2021 | Effect of horticultural therapy on apathy in nursing home residents with dementia: a pilot randomized controlled trial | Not a nature intervention |
| Jin_2015 | Effect of outdoor activity on myopia onset and progression in school-aged children in northeast China: the Sujiatun Eye Care Study. | Not a nature intervention |
| NCT03304301_2017 | Effect of Sunlight Exposure and Outdoor Activities on Depression, Cognition and Quality of Life in the Elderly | Protocol without published results |
| He_2015 | Effect of Time Spent Outdoors at School on the Development of Myopia Among Children in China: A Randomized Clinical Trial. | School programmes with outdoor play, no clear nature focus |
| NCT04958499_2021 | Effectiveness of Bio-Healthy Park on Adult | Ongoing study |
| Chang_2016 | Effectiveness of community-based exercise intervention programme in obese adults with metabolic syndrome. | Not a nature intervention |
| Elley_2003 | Effectiveness of counselling patients on physical activity in general practice: cluster randomised controlled trial | Not a nature intervention |
| Gatzemann_2008 | Effectiveness of sports activities with an orientation on experiential education, adventure-based learning and outdoor-education | Complex outdoor programme |
| Jung_2018 | Effectiveness of the KENKOJISEICHI local revitalization system on cognitive function change in older adults with mild cognitive impairment: study protocol for a randomized controlled trial. | Protocol without published results |
| Lin_2020 | Effects of a combination of three-dimensional virtual reality and hands-on horticultural therapy on institutionalized older adults physical and mental health: quasi-experimental design | Digital representation of nature |
| Lin_2020 | Effects of a combination of three-dimensional virtual reality and hands-on horticultural therapy on institutionalized older adults[left right double arrow] physical and mental health: quasi-experimental design | Duplicate / already included |
| Schofield_2005 | Effects of a controlled pedometer-intervention trial for low-active adolescent girls. | Not a nature intervention |
| Suemaru_1996 | Effects of a long term trial of Fukuyama Yuai-Outdoor Activity Care Programme (FY-OACP) on behavioural disorders, hypothalamic-pitutary-adrenal axis and immune activity in demented elderly patients | Wrong publication type |
| Modesto_2021 | Effects of a Real-Life Park-Based Physical Activity Interventional Program on Cardiovascular Risk and Physical Fitness | Single-group design |
| Saakslahti_2004 | Effects of a three-year intervention on children's physical activity from age 4 to 7. | Not a nature intervention |
| Wu_2015 | Effects of an 8-Week Outdoor Brisk Walking Program on Fatigue in Hi-Tech Industry Employees. | Not a nature intervention |
| Wu_2015 | Effects of an 8-Week Outdoor Brisk Walking Program on Fatigue in Hi-Tech Industry Employees: a Randomized Control Trial | Duplicate / already included |
| Siltanen_2020 | Effects of an Individualized Active Aging Counseling Intervention on Mobility and Physical Activity: secondary Analyses of a Randomized Controlled Trial | Not a nature intervention |
| Gustafsson_2012 | Effects of an outdoor education intervention on the mental health of schoolchildren. | School programmes with outdoor play, no clear nature focus |
| Song_2020 | Effects of an Urban Forest-Based Health Promotion Program on Children Living in Group Homes. | Single-group design |
| NCT04903171_2021 | Effects of Enriched Gardens in Nursing Home Residents With Dementia | Ongoing study |
| Makizako_2015 | Effects of exercise and horticultural intervention on the brain and mental health in older adults with depressive symptoms and memory problems: study protocol for a randomized controlled trial [UMIN000018547]. | Duplicate / already included |
| Wu_2017 | Effects of forest bathing on plasma endothelin-1 in elderly patients with chronic heart failure: Implications for adjunctive therapy | Wrong publication type |
| JPRN-UMIN000040588_2020 | Effects of group walking in forests and hills on physical and mental functions of community-dwelling elderly | Ongoing study |
| Chen_2015 | Effects of Horticultural Therapy on Psychosocial Health in Older Nursing Home Residents: A Preliminary Study. | Single-group design |
| Oh_2012 | Effects of Horticultural Therapy Program on State-Anxiety, Fatigue and Quality of Life among Women Cancer Survivors. | Not in English |
| LaiCKY_2018 | Effects of Horticulture on Frail and Prefrail Nursing Home Residents: a Randomized Controlled Trial | Not a nature intervention |
| Yao_2017 | Effects of horticulture therapy on nursing home older adults in southern Taiwan | Not a nature intervention |
| Yao_2016 | Effects of horticulture therapy on nursing home older adults in southern Taiwan. | Duplicate / already included |
| Kang_2021 | Effects of integrated indirect forest experience on emotion, fatigue, stress and immune function in hemodialysis patients | Digital representation of nature |
| NCT04781491_2021 | Effects of Nature and Forest Therapy in Patients With Metabolic Syndrome and Cardiovascular Risk Factors | Ongoing study |
| NCT03716440_2018 | Effects of Nature Exposure on Smoking Behavior | Protocol without published results |
| Nagyova_2020 | Effects of Nordic walking on cardiovascular performance and quality of life in coronary artery disease | Not a nature intervention |
| NAGYOVA_2020 | Effects of Nordic walking on cardiovascular performance and quality of life in coronary artery disease | Not a nature intervention |
| Vahlberg_2016 | Effects of SMS-guided outdoor walking and strength training after acute stroke-a pilot study | Not a nature intervention |
| Wolfenden_2019 | Efficacy of a free-play intervention to increase physical activity during childcare: a randomized controlled trial. | School programmes with outdoor play, no clear nature focus |
| NCT04291183_2020 | Elderly Who Received Horticulturel Therapy | Ongoing study |
| Song_2015 | Elucidation of a physiological adjustment effect in a forest environment: a pilot study | Wrong publication type |
| Song_2014 | Elucidation of the physiological adjustment effect of forest therapy | Not in English |
| Han_2018 | Empirical examinations of effects of three-level green exercise on engagement with nature and physical activity | Not an interventional study |
| JPRN-UMIN000011398_2013 | Empirical study of the horticultural therapy as a means of the local community reproduction support in a disaster area | Protocol without published results |
| Sianoja_2018 | Enhancing daily well-being at work through lunchtime park walks and relaxation exercises: recovery experiences as mediators | No statistical comparison |
| Bourdon_2021 | Enriched gardens improve cognition and independence of nursing home residents with dementia: a pilot controlled trial | Inappropriate comparison |
| Benton_2018 | Evaluating the impact of improvements in urban green space on older adults' physical activity and wellbeing: protocol for a natural experimental study. | Not a behavioural change intervention |
| KamMCY_2010 | Evaluation of a horticultural activity programme for persons with psychiatric illness | Duplicate / already included |
| ISRCTN59395217_2020 | Examining the effects of outdoor recreational experiences on military veterans with PTSD | Ongoing study |
| NCT00958867_2009 | EXCEL: exercise for Cognition and Everyday Living for Seniors With Memory Complaints | Not a nature intervention |
| ACTRN12620000733976_2020 | Exercise interveNtion outdoor proJect in the cOmmunitY (ENJOY) program in Dementia | Ongoing study |
| Levinger_2020 | Exercise interveNtion outdoor proJect in the cOmmunitY for older people - results from the ENJOY Seniors Exercise Park project translation research in the community. | Single-group design |
| Levinger_2019 | Exercise interveNtion outdoor proJect in the cOmmunitY for older people - the ENJOY Senior Exercise Park project translation research protocol. | Single-group design |
| Nost_2016 | Expectations, effect and experiences of an easily accessible self-management intervention for people with chronic pain: study protocol for a randomised controlled trial with embedded qualitative study | Not a nature intervention |
| Wolf_2011 | Experiential learning in psychotherapy: ropes course exposures as an adjunct to inpatient treatment. | Complex outdoor programme |
| Williams_2017 | Face-to-face versus telephone delivery of the green prescription for Maori and New Zealand Europeans with type-2 diabetes mellitus: influence on participation and health outcomes | Inappropriate comparison |
| Beavers_2017 | Feasibility of a pilot randomized controlled trial to assess health impacts of community gardening | Wrong publication type |
| Yang_2019 | Feasibility of an outdoor mindful walking program for reducing negative affect in older adults | Not a nature intervention |
| Yang_2018 | Feasibility of an Outdoor Mindful Walking Program for Reducing Negative Affect in Older Adults. | Duplicate / already included |
| NCT04878744_2021 | Feasibility Study of an App-based Intervention to Improve Wellbeing | Ongoing study |
| Spees_2016 | Feasibility, Preliminary Efficacy, and Lessons Learned From a Garden-Based Lifestyle Intervention for Cancer Survivors. | Single-group design |
| Farmer_1995 | Field trips and follow-up activities: Fourth graders in a public garden. | No appropriate outcomes measured |
| Anderson_2011 | Findings from a Pilot Investigation of the Effectiveness of a Snoezelen Room in Residential Care: Should We Be Engaging with Our Residents More? | No data given for nature intervention |
| Uhls_2014 | Five days at outdoor education camp without screens improves preteen skills with nonverbal emotion cues. | Complex outdoor programme |
| Kim_2015 | Forest adjuvant anti-cancer therapy to enhance natural cytotoxicity in urban women with breast cancer: A preliminary prospective interventional study | Single-group design |
| Peterfalvi_2021 | Forest bathing always makes sense: Blood pressure-lowering and immune system-balancing effects in late spring and winter in central europe | Single-group design |
| Machackova_2021 | Forest manners exchange: Forest as a place to remedy risky behaviour of adolescents: Mixed methods approach | No health outcomes measured |
| Kim_2021 | Forest therapy alone or with a guide: Is there a difference between self-guided forest therapy and guided forest therapy programs? | Inappropriate comparison |
| Warber_2021 | Fostering holistic and spiritual wellbeing for older women: a protocol for a usbased feasibility study of forest bathing | Ongoing study |
| Caramia_2017 | Gait parameters are differently affected by concurrent smartphone-based activities with scaled levels of cognitive effort | Not a nature intervention |
| Stluka_2019 | Gardening for health: Using garden coordinators and volunteers to implement Rural School and community gardens | No appropriate outcomes measured |
| Cases_2015 | Gardening intervention increases telomerase levels in breast cancer survivors | Wrong publication type |
| Barclay_2018 | Getting older adults OUTdoors (GO-OUT) | Duplicate / already included |
| NCT02339467_2015 | Getting Older Adults OUT-of -Doors | Duplicate / already included |
| ISRCTN99926592_2018 | Green exercise and bathing therapy for the treatment of non-specific chronic low back pain | Complex outdoor programme |
| Huber_2019 | Green exercise and mg-ca-SO4 thermal balneotherapy for the treatment of non-specific chronic low back pain: a randomized controlled clinical trial | Complex outdoor programme |
| Irvine_2020 | Group outdoor health walks using activity trackers: Measurement and implementation insight from a mixed-methods feasibility study | Wrong publication type |
| Blair_2013 | Harvest for health gardening intervention feasibility study in cancer survivors | Single-group design |
| Gigliotti_2004 | Harvesting health: effects of three types of horticultural therapy activities for persons with dementia. | Inappropriate comparison |
| Zick_2013 | Harvesting More Than Vegetables: The Potential Weight Control Benefits of Community Gardening. | Not an interventional study |
| Razani_2015 | Healing through Nature: A Park-Based Health Intervention for Young People in Oakland, California | Qualitative study |
| NCT02094144_2014 | Health Benefits of a 6-month Brisk Walking Program in Sedentary Postmenopausal Women | Not a nature intervention |
| NCT03266120_2017 | Health Benefits of Gardening | Protocol without published results |
| Jia_2016 | Health Effect of Forest Bathing Trip on Elderly Patients with Chronic Obstructive Pulmonary Disease | Wrong publication type |
| Kolt_2011 | Healthy steps trial: effectiveness of a pedometer-based green prescription for low-active older adults in primary care | Not a nature intervention |
| Kolt_2012 | Healthy Steps trial: pedometer-based advice and physical activity for low-active older adults | Not a nature intervention |
| Pope_2019 | Heart rate variability differences between green and suburban walking: a pilot crossover study | Wrong publication type |
| Lunt_2014 | High intensity interval training in a real world setting: a randomized controlled feasibility study in overweight inactive adults, measuring change in maximal oxygen uptake | Inappropriate comparison |
| Kotozaki_2014 | Horticultural therapy as a measure for recovery support of regional community in the disaster area: a preliminary experiment for forty five women who living certain region in the coastal area of Miyagi Prefecture | Not in English |
| IRCT20170611034454N3_2018 | horticultural therapy on quality of life | Protocol without published results |
| Oh_2020 | Horticultural therapy program for improving emotional well-being of elementary school students: an observational study. | Single-group design |
| Wong_2020 | Horticultural Therapy Reduces Biomarkers of Immunosenescence and Inflammaging in Community-Dwelling Older Adults: a Feasibility Pilot Randomized Controlled Trial | Duplicate / already included |
| Detweiler_2015 | Horticultural therapy: a pilot study on modulating cortisol levels and indices of substance craving, posttraumatic stress disorder, depression, and quality of life in veterans | Duplicate / already included |
| Beela_2015 | Horticulture Therapy for the Improvement of Self Concept in Adolescents with Locomotor and Hearing Impairment. | Single-group design |
| NCT02341716_2015 | Hospital- and Home-based Supervised Exercise Versus UNsupervised Walk Advice For Patients With InTermittent Claudication | Ongoing study |
| Verra_2015 | How much does therapeutic nordic walking help people to cope with psychosomatic disorders? | Not a nature intervention |
| Morrier_2018 | I Wanna Play Too: Factors Related to Changes in Social Behavior for Children With and Without Autism Spectrum Disorder After Implementation of a Structured Outdoor Play Curriculum. | School programmes with outdoor play, no clear nature focus |
| Cohen_2012 | Impact and cost-effectiveness of family Fitness Zones: a natural experiment in urban public parks. | Not a behavioural change intervention |
| Carney_2012 | Impact of a Community Gardening Project on Vegetable Intake, Food Security and Family Relationships: A Community-based Participatory Research Study. | No appropriate outcomes measured |
| Sharpe_2010 | Impact of a community-based prevention marketing intervention to promote physical activity among middle-aged women. | Not a nature intervention |
| Toews_2018 | Impact of a nature-based intervention on incarcerated women | Single-group design |
| Spees_2019 | Impact of a Tailored Nutrition and Lifestyle Intervention for Overweight Cancer Survivors on Dietary Patterns, Physical Activity, Quality of Life, and Cardiometabolic Profiles. | Single-group design |
| Sobko_2020 | Impact of outdoor nature-related activities on gut microbiota, fecal serotonin, and perceived stress in preschool children: the Play&Grow randomized controlled trial | No appropriate outcomes measured |
| Razak_2018 | Impact of scheduling multiple outdoor free-play periods in childcare on child moderate-to-vigorous physical activity: a cluster randomised trial | School programmes with outdoor play, no clear nature focus |
| Tucker_2017 | Impact of the Supporting Physical Activity in the Childcare Environment (SPACE) intervention on preschoolers' physical activity levels and sedentary time: a single-blind cluster randomized controlled trial | Not a nature intervention |
| Langhammer_2010 | Improving gait after stroke - Treadmill or walking outdoors | Not a nature intervention |
| McCluskey_2013 | Improving quality of life by increasing outings after stroke: study protocol for the Out-and-About trial | Not a nature intervention |
| Stanley_2016 | Increasing physical activity among young children from disadvantaged communities: study protocol of a group randomised controlled effectiveness trial | Not a nature intervention |
| Brown_2004 | Indoor gardening older adults: effects on socialization, activities of daily living, and loneliness | Inappropriate comparison |
| JuyoungLee_2014 | Influence of Forest Therapy on Cardiovascular Relaxation in Young Adults. | Single-group design |
| Weaver_2018 | Initial Outcomes of a Participatory-Based, Competency-Building Approach to Increasing Physical Education Teachersâ€™ Physical Activity Promotion and Studentsâ€™ Physical Activity: A Pilot Study. | Not a nature intervention |
| NCT02573142_2015 | Integrated Child Obesity Treatment Study: bull City Healthy and Fit | Duplicate / already included |
| Jansson_2019 | Integrating smartphone technology, social support and the outdoor built environment to promote community-based aerobic and resistance-based physical activity: Rationale and study protocol for the 'ecofit' randomized controlled trial | Duplicate / already included |
| Jansson_2019 | Integrating smartphone technology, social support and the outdoor built environment to promote community-based aerobic and resistance-based physical activity: Rationale and study protocol for the 'ecofit' randomized controlled trial | Duplicate / already included |
| Plotnikoff_2017 | Integrating smartphone technology, social support and the outdoor physical environment to improve fitness among adults at risk of, or diagnosed with, Type 2 Diabetes: findings from the 'eCoFit' randomized controlled trial | Duplicate / already included |
| NCT02124837_2014 | Intervention Study on Break Activities and Workers[spacing acute] Psychological and Physiological Health & Performance | Duplicate / already included |
| Kerse_2005 | Is physical activity counseling effective for older people? A cluster randomized, controlled trial in primary care | Not a nature intervention |
| Mitchell_2013 | Is physical activity in natural environments better for mental health than physical activity in other environments? | Not an interventional study |
| Inouye_2014 | Lifestyle Intervention for Filipino Americans at Risk for Diabetes. | Not a nature intervention |
| NCT00425269_2007 | Lifestyle Intervention for Pakistani Women in Oslo | Protocol without published results |
| Nowak_2014 | Live music promotes positive behaviours in people with Alzheimer's disease | Not a nature intervention |
| Slining_2021 | LiveWell in early childhood: results from a two-year pilot intervention to improve nutrition and physical activity policies, systems and environments among early childhood education programs in South Carolina. | School programmes with outdoor play, no clear nature focus |
| Arbillaga-Etxarri_2018 | Long-term efficacy and effectiveness of a behavioural and community-based exercise intervention (Urban Training) to increase physical activity in patients with COPD: a randomised controlled trial | Not a behavioural change intervention |
| Ottosson_2005 | Measures of restoration in geriatric care residences: The influence of nature on elderly people's power of concentration, blood pressure and pulse rate | Not an interventional study |
| Wagenfeld_2019 | Measuring Emotional Response to a Planting Activity for Staff at an Urban Office Setting: A Pilot Study. | Single-group design |
| vanStralen_2012 | Mediators of the effect of the JUMP-in intervention on physical activity and sedentary behavior in Dutch primary schoolchildren from disadvantaged neighborhoods. | Not a nature intervention |
| Farrier_2019 | Mental health and wellbeing benefits from a prisons horticultural programme. | Qualitative study |
| DRKS00025230_2021 | Mobility in Old age By Integrating health care and personal network resources in older adults Living in rural arEas | Protocol without published results |
| Matsouka_2005 ;_ | Mood alterations following an indoor and outdoor exercise program in healthy elderly women | Not a nature intervention |
| Tortella_2016 | Motor skill development in Italian pre-school children induced by structured activities in a specific playground | School programmes with outdoor play, no clear nature focus |
| ISRCTN43292449_2020 | Mountain hiking versus nature connection therapy based in Algund as climate therapy for couples | Protocol without published results |
| JPRN-UMIN000036888_2019 | Multi-component cognitive intervention with physical exercise for older adults with Alzheimer's disease: a 6-month randomized controlled trial | Protocol without published results |
| NCT01885325_2013 | Multi-component Intervention to Increase Physical Activity in Preschool Children | School programmes with outdoor play, no clear nature focus |
| Santos_2021 | Multicomponent physical activity program to prevent body changes and metabolic disturbances associated with antiretroviral therapy and improve quality of life of people living with HIV: a pragmatic trial | Not a nature intervention |
| Wong-Yu_2015 | Multi-dimensional balance training programme improves balance and gait performance in people with Parkinson's disease: a pragmatic randomized controlled trial with 12-month follow-up | Not a nature intervention |
| Li_2005 | Multilevel modelling of built environment characteristics related to neighbourhood walking activity in older adults. | Not a behavioural change intervention |
| Wu_2018 | Myopia Prevention and Outdoor Light Intensity in a School-Based Cluster Randomized Trial | School programmes with outdoor play, no clear nature focus |
| Marselle_2012 | Natural health service: Enhancing wellbeing with group walks in green spaces | Wrong publication type |
| Shrestha_2021 | Natural or Urban Campus Walks and Vitality in University Students: exploratory Qualitative Findings from a Pilot Randomised Controlled Study | Qualitative study |
| VarningPoulsen_2020 | Nature is just around us! Development of an educational program for implementation of nature-based activities at a crisis shelter for women and children exposed to domestic violence | Protocol without published results |
| Bai_2020 | Nature play and fundamental movement skills training programs improve childcare educator supportive physical activity behavior | No health outcomes measured |
| Sia_2020 | Nature-based activities improve the well-being of older adults | Single-group design |
| NCT04897685_2021 | Nature-based Treatment Group for Depression | Ongoing study |
| Granziera_2021 | Nordic Walking and Walking in Parkinson's disease: a randomized single-blind controlled trial | Not a nature intervention |
| Breyer_2010 | Nordic walking improves daily physical activities in COPD: a randomised controlled trial | Not a nature intervention |
| ISRCTN31525632_2009 | Nordic walking to improve functional exercise capacity and daily physical activities in chronic obstructive pulmonary disease (COPD) | Not a nature intervention |
| Passmore_2017 | Noticing nature: Individual and social benefits of a two-week intervention. | No appropriate outcomes measured |
| Braun_2019 | NP23 Garden-Based Intervention for Youth Improves Dietary and Physical Activity Patterns, Quality of Life, Family Relationships, and Indices of Health...Society for Nutrition Education and Behavior 52nd Annual Conference, Nutrition Education: Rooted in Fo | Wrong publication type |
| Lee_2021 | Nurtured in Nature: a Pilot Randomized Controlled Trial to Increase Time in Greenspace among Urban-Dwelling Pregnant Women | Duplicate / already included |
| Wooller_2016 | Occlusion of sight, sound and smell during Green Exercise influences mood, perceived exertion and heart rate | Inappropriate comparison |
| Mori_2021 | Occupational Therapy and Therapeutic Horticulture for Women with Cancer and Chronic Pain: A Pilot Study. | Single-group design |
| Patel_2020 | Older adults' evaluations of the standard and modified pedometer-based Green Prescription | Not a nature intervention |
| DRKS00015117_2018 | OUTDOOR ACTIVE - Development of a community-based outdoor physical activity promotion program in older adults 65+ | Protocol without published results |
| McAvoy_2006 | Outdoor adventure programming for individuals with cognitive disabilities who present serious accommodation challenges. | Complex outdoor programme |
| Gill_2016 | Outdoor adventure therapy to increase physical activity in young adult cancer survivors. | Complex outdoor programme |
| Wheeler_2020 | Outdoor recreational activity experiences improve psychological wellbeing of military veterans with post-traumatic stress disorder: Positive findings from a pilot study and a randomised controlled trial | Complex outdoor programme |
| Langhammer_2010 | Outdoors or indoors walking, what is more beneficial? A comparison of exercise methods in a randomized trial | Not a nature intervention |
| Zarr_2017 | Park Prescription (DC Park Rx): A New Strategy to Combat Chronic Disease in Children | Wrong publication type |
| NCT04114734_2019 | Park Rx and Physical Activity Among Low-income Children | Ongoing study |
| Bush_2007 | Park-based obesity intervention program for inner-city minority children. | Single-group design |
| NCT02337075_2015 | PAthway To Health (PATH): fostering Culturally Appropriate Physical Activities in Indigenous Communities | Protocol without published results |
| Christiana_2016 | Pediatrician prescriptions for outdoor physical activity among children: A pilot study | Not a nature intervention |
| Patel_2013 | Perceived barriers, benefits, and motives for physical activity: Two primary-care physical activity prescription programs | Not a nature intervention |
| Kien_2003 | Physical activity in middle school-aged children participating in a school-based recreation program | School programmes with outdoor play, no clear nature focus |
| Cohen_2013 | Physical activity in parks: A randomized controlled trial using community engagement. | Not a behavioural change intervention |
| Shvedko_2019 | Physical activity intervention for loneliness (PAIL) in community-dwelling older adults: a randomised feasibility study | Not a nature intervention |
| Pasek_2020 | Physical fitness as part of the health and well-being of students participating in physical education lessons indoors and outdoors | School programmes with outdoor play, no clear nature focus |
| ISRCTN14058106_2017 | Physical Literacy in the Early Years (PLEY) project : a loose parts intervention to promote active outdoor play in preschool aged children | School programmes with outdoor play, no clear nature focus |
| Dasgupta_2017 | Physician step prescription and monitoring to improve ARTERial health (SMARTER): a randomized controlled trial in patients with type 2 diabetes and hypertension | Not a nature intervention |
| Joseph_2017 | Physician step prescription and monitoring to improve ARTERial health (SMARTER): A randomized controlled trial in patients with type 2 diabetes and hypertension | Not a nature intervention |
| Dasgupta_2017 | Physician step prescription and monitoring to improve ARTERial health (SMARTER): a randomized controlled trial in patients with type 2 diabetes and hypertension | Duplicate / already included |
| Ochiai_2015 | Physiological and psychological effects of a forest therapy program on middle-aged females | Single-group design |
| Ikei_2014 | Physiological and psychological effects of viewing forest landscapes in a seated position in one-day forest therapy experimental model | Not in English |
| Lee_2011 | Physiological benefits of forest environment: based on field research at 4 sites | Not in English |
| NCT04654949_2020 | Pilot Randomized Controlled Trial: horticultural Therapy for Inpatient Older Adults in an Acute Care Hospital | Ongoing study |
| Olafsdottir-2017 | Place, green exercise and stress: an exploration of lived experience and restorative effects | Qualitative study |
| Aaron_2010 | Planting a seed: An examination of nature perception, program processes, and outdoor experience. | Wrong publication type |
| Scott_2020 | Positive aging benefits of home and community gardening activities: Older adults report enhanced self-esteem, productive endeavours, social engagement and exercise | Not an interventional study |
| Kondo_2011 | Positive healthy physiological effects of Shinrin-yoku in human. | Not in English |
| Chen_2020 | Potential benefits of environmental volunteering programs of the health of older adults: a pilot study. | Single-group design |
| Yoon_2016 | Preliminary Effectiveness and Sustainability of Group Aerobic Exercise Program in Patients with Schizophrenia. | Not a nature intervention |
| James_2016 | Prescribing Outdoor Play: Outdoors Rx | No statistical comparison |
| NCT04917666_2021 | Process and Outcomes of Horticultural Therapy for People With Disabilities | Ongoing study |
| Burkow_2018 | Promoting exercise training and physical activity in daily life: a feasibility study of a virtual group intervention for behaviour change in COPD. | Not a nature intervention |
| Sobko_2017 | Promoting healthy eating and active playtime by connecting to nature families with preschool children: evaluation of pilot study "play&Grow" | No appropriate outcomes measured |
| Sobko_2016 | Promoting healthy eating and active playtime by connecting to nature families with preschool children: evaluation of pilot study "Play&Grow". | Duplicate / already included |
| NCT04329052_2020 | Promoting Mental Well-being for Secondary School Students Through an Experiential Learning Activity | Complex outdoor programme |
| Stubbs_2002 | Promoting participation in physical activity in a community intervention study. | No appropriate outcomes measured |
| IRCT20191011045056N1_2020 | Promoting self-care, health literacy and social capital in older adults | Protocol without published results |
| Balcazar_2015 | Promotoras Can Facilitate Use of Recreational Community Resources: The Mi Corazon Mi Comunidad Cohort Study | No health outcomes measured |
| Vahlberg_2018 | Protocol and pilot study of a short message service-guided training after acute stroke/transient ischemic attack to increase walking capacity and physical activity | Not a nature intervention |
| Ourania_2012 | Psychological and Physiological Effects of Aquatic Exercise Program Among the Elderly. | Wrong publication type |
| Morita_2007 | Psychological effects of forest environments on healthy adults: Shinrin-yoku (forest-air bathing, walking) as a possible method of stress reduction | Single-group design |
| Sellman_2017 | Psychosocial enhancement of the Green Prescription for obesity recovery: a randomised controlled trial | Not a nature intervention |
| Bassi_2018 | Quality of experience during horticultural activities: an experience sampling pilot study among older adults living in a nursing home | No health outcomes measured |
| Logan_2004 | Randomised controlled trial of an occupational therapy intervention to increase outdoor mobility after stroke | Not a nature intervention |
| JPRN-UMIN000009260_2012 | Randomized controlled trial of integrative medicine program for self management on fibromyalgia | Protocol without published results |
| Litt_2018 | Rationale and design for the community activation for prevention study (CAPs): a randomized controlled trial of community gardening | Protocol without published results |
| Armstrong_2020 | Rationale and design of "Hearts & Parks": study protocol for a pragmatic randomized clinical trial of an integrated clinic-community intervention to treat pediatric obesity. | Not a nature intervention |
| Cortinez-O'Ryan_2017 | Reclaiming streets for outdoor play: A process and impact evaluation of "Juega en tu Barrio" (Play in your Neighborhood), an intervention to increase physical activity and opportunities for play | Not a nature intervention |
| White_2016 | Recreational physical activity in natural environments and implications for health: A population based cross-sectional study in England | Not an interventional study |
| Messiah_2016 | Reducing childhood obesity through coordinated care: Development of a park prescription program | Protocol without published results |
| Logan_2014 | Rehabilitation aimed at improving outdoor mobility for people after stroke: a multicentre randomized controlled study (the getting out of the house study) | Not a nature intervention |
| Kang_2015 | Relief of Chronic Posterior Neck Pain Depending on the Type of Forest Therapy: comparison of the Therapeutic Effect of Forest Bathing Alone Versus Forest Bathing With Exercise | Inappropriate comparison |
| Pasanen_2018 | Restoration, well-being, and everyday physical activity in indoor, built outdoor and natural outdoor settings. | Not an interventional study |
| Barclay_2018 | Safety and feasibility of an interactive workshop and facilitated outdoor walking group compared to a workshop alone in increasing outdoor walking activity among older adults: a pilot randomized controlled trial | No statistical comparison |
| Wells_2014 | School gardens and physical activity: a randomized controlled trial of low-income elementary schools | Duplicate / already included |
| Wells_2014 | School gardens and physical activity: a randomized controlled trial of low-income elementary schools. | School programmes with outdoor play, no clear nature focus |
| Guijarro-Romero_2021 | School physical education-based reinforced program through moderate-to-vigorous physical activity improves and maintains schoolchildren's cardiorespiratory fitness: A cluster-randomized controlled trial | School programmes with outdoor play, no clear nature focus |
| Sinclair_2007 | Self-reported health benefits in patients recruited into New Zealand's 'Green Prescription' primary health care program | Not a nature intervention |
| Lopez-Pousa_2015 | Sense of Well-Being in Patients with Fibromyalgia: aerobic Exercise Program in a Mature Forest - A Pilot Study | Inappropriate comparison |
| NCT04139421_2019 | Shinrin-Yoku (Forest Bathing) on Psychological Well-being | Ongoing study |
| Nost_2018 | Short-term effect of a chronic pain self-management intervention delivered by an easily accessible primary healthcare service: a randomised controlled trial | Not a nature intervention |
| Lin_2017 | Short-Term Efficacy of a "Sit Less, Walk More" Workplace Intervention on Improving Cardiometabolic Health and Work Productivity in Office Workers. | Not a nature intervention |
| NCT04328363_2020 | Social Prescription and Lifestyles Modification to Reduce Glycemia in People With Prediabetes (PREDIBAL) | Ongoing study |
| Cassarino_2019 | Sometimes Nature Doesn't Work: Absence of Attention Restoration in Older Adults Exposed to Environmental Scenes. | Digital representation of nature |
| Shigematsu_2008 | Square-stepping exercise and fall risk factors in older adults: a single-blind, randomized controlled trial | Not a nature intervention |
| Daskalopoulou_2014 | Step Monitoring to improve ARTERial health (SMARTER) through step count prescription in type 2 diabetes and hypertension: Trial design and methods | Not a nature intervention |
| Dasgupta_2014 | Step Monitoring to improve ARTERial health (SMARTER) through step count prescription in type 2 diabetes and hypertension: trial design and methods | Duplicate / already included |
| ACTRN12620001183976_2020 | Stepped Wedge Cluster Randomised Trial of Social Prescribing of Forest Therapy for Adults with Mental Illness | Ongoing study |
| Thomas_2020 | Stepped-Wedge Cluster Randomised Trial of Social Prescribing of Forest Therapy for Quality of Life and Biopsychosocial Wellbeing in Community-Living Australian Adults with Mental Illness: protocol | Ongoing study |
| Widodo_2019 | Stress of brain mapping in elderly people before and after giving horticultural therapy in planting flowers | No appropriate outcomes measured |
| NCT04314388_2020 | Stroke in Young Adults Outdoor Rehabilitation | Ongoing study |
| Stolley_2015 | Study design and protocol for moving forward: a weight loss intervention trial for African-American breast cancer survivors | Not a nature intervention |
| Pearson_2020 | Study of active neighborhoods in Detroit (StAND): study protocol for a natural experiment evaluating the health benefits of ecological restoration of parks. | Not a behavioural change intervention |
| Egawa_2003 | Study on efficient intervention frequency of functional fitness promotion program for community-dwelling elderly people part2: Effect of intervention frequency on physical, mental, and social functional capacity | Not a nature intervention |
| Wells_2014 | Study protocol: effects of school gardens on children's physical activity. | Protocol without published results |
| Vanderloo_2016 | Supporting Physical Activity in the Childcare Environment (SPACE): A Cluster Randomized Controlled Trial...2016 North American Society for Pediatric Exercise Medicine (NASPEM) Biennial Meeting, Knoxville, Tennessee. | Not a nature intervention |
| Tucker_2016 | Supporting Physical Activity in the Childcare Environment (SPACE): rationale and study protocol for a cluster randomized controlled trial. | Not a nature intervention |
| Wong-Yu_2015 | Task- and Context-Specific Balance Training Program Enhances Dynamic Balance and Functional Performance in Parkinsonian Nonfallers: A Randomized Controlled Trial With Six-Month Follow-Up. Arch Phys Med Rehabil. | Not a nature intervention |
| ISRCTN56739489_2014 | The Blossom Project: moms2Move (M2M) | Protocol without published results |
| Wiersma_2012 | The Development and Pilot Testing of Active Kids: A Park-Based Afterschool Physical Activity Program for Hispanic Youth. | Single-group design |
| Mackay_2010 | The effect of "green exercise" on state anxiety and the role of exercise duration, intensity, and greenness: A quasi-experimental study | Single-group design |
| Weltin_2012 | The Effect of a Community Garden on HgA1c in Diabetics of Marshallese Descent. | Inappropriate comparison |
| Rantanen_2015 | The effect of an outdoor activities' intervention delivered by older volunteers on the quality of life of older people with severe mobility limitations: a randomized controlled trial | Not a nature intervention |
| Rantanen_2014 ;_ | The effect of an outdoor activities' intervention delivered by older volunteers on the quality of life of older people with severe mobility limitations: a randomized controlled trial. | Duplicate / already included |
| Duncan_2014 | The effect of green exercise on blood pressure, heart rate and mood state in primary school children | Digital representation of nature |
| Rantakokko_2015 | The effect of out-of-home activity intervention delivered by volunteers on depressive symptoms among older people with severe mobility limitations: a randomized controlled trial. | Not a nature intervention |
| IRCT20190207042649N1_2019 | The Effect of Physical Activity in Outdoor and Indoor Space on Motor, Perceptual and Social Development among Preschool Children | Protocol without published results |
| Palmer_2019 | The effect of the CHAMP intervention on fundamental motor skills and outdoor physical activity in preschoolers | School programmes with outdoor play, no clear nature focus |
| Marcy-Edwards_2011 | The effect of vignette activity on the neuropsychiatric behaviours expressed by individuals with dementia, living in long-term care | Wrong publication type |
| Irandoust_2017 | The Effect of Vitamin D supplement and Indoor Vs Outdoor Physical Activity on Depression of Obese Depressed Women. | Not a nature intervention |
| Choi_2014 | The Effectiveness of a Forest-experience-integration Intervention for Community Dwelling Cancer Patients' Depression and Resilience. | Not in English |
| NTR2483_2010 | The effectiveness of an internet physical activity/self-management program in patients with osteoarthritis who did not recently receive a treatment | Not a nature intervention |
| Zachor_2017 | The effectiveness of an outdoor adventure programme for young children with autism spectrum disorder: a controlled study. | Complex outdoor programme |
| DRKS00016370_2019 | The effectiveness of smartphone app guided interval-walking training compared to continuous aerobic walking training for people with type 2 diabetes or prediabetes. A pilot study | Protocol without published results |
| Banda_2017 | The Effects of a Park Awareness Campaign on Rural Park Use and Physical Activity. | Unit of observation is parks, not people |
| Tesler_2018 | The effects of an urban forest health intervention program on physical activity, substance abuse, psychosomatic symptoms, and life satisfaction among adolescents | Complex outdoor programme |
| Pirchio_2021 | The Effects of Contact With Nature During Outdoor Environmental Education on Students' Wellbeing, Connectedness to Nature and Pro-sociality. | School programmes with outdoor play, no clear nature focus |
| Yamaguchi_2006 | The effects of exercise in forest and urban environments on sympathetic nervous activity of normal young adults | No appropriate outcomes measured |
| White_2015 | The effects of exercising in different natural environments on psycho-physiological outcomes in post-menopausal women: A simulation study | Digital representation of nature |
| NgKST_2016 | The effects of horticultural therapy on the psychological well-being and associated biomarkers of elderly in Singapore | Wrong publication type |
| NgKST_2017 | The effects of horticultural therapy on the psychoneuroimmunological markers of elderly in Singapore: second wave findings from the randomized controlled trial | Wrong publication type |
| Alhassan_2007 | The effects of increasing outdoor play time on physical activity in Latino preschool children | Not a nature intervention |
| Kim_2020 | The effects of outdoor activities in forests on atopic dermatitis | Wrong publication type |
| Prins_2019 | The effects of small-scale physical and social environmental interventions on walking behaviour among Dutch older adults living in deprived neighbourhoods: results from the quasi-experimental NEW.ROADS study. | Not a behavioural change intervention |
| ISRCTN66167429_2017 | The effects of trail versus road running on neuromuscular performance | Not a nature intervention |
| Bang_2016 | The Effects of Urban Forest-walking Program on Health Promotion Behavior, Physical Health, Depression, and Quality of Life: a Randomized Controlled Trial of Office-workers | Not in English |
| Buru_2019 | The evaluation of therapeutic horticulture effects on urinary tryptophan metabolites by using the spectrofluorimetric analysis in a group of students | Wrong publication type |
| Swinburn_1998 | The green prescription study: a randomized controlled trial of written exercise advice provided by general practitioners | Not a nature intervention |
| Wood_2020 | The health impact of nature exposure and green exercise across the life course: a pilot study | Not an interventional study |
| Kolt_2009 | The healthy steps study: a randomized controlled trial of a pedometer-based green prescription for older adults. Trial protocol | Not a nature intervention |
| N/A | The impact of an exercise program based on nordic walking on patients with a recent acute coronary syndrome. | Not a nature intervention |
| ISRCTN81041724_2006 | The impact of implementation intentions in changing complex health-related behaviours in order to prevent weight gain: the case of physical activity | Not a nature intervention |
| Wade_2019 | The influence of exercise in the natural environment on adolescents’ sustained attention and working memory: A randomised controlled trial | Wrong publication type |
| Shin_2011 | The influence of forest therapy camp on depression in alcoholics | Duplicate / already included |
| Shin_2011 | The influence of forest therapy camp on depression in alcoholics | Complex outdoor programme |
| Toda_2015 | The influence of personal patterns of behavior on the physiological effects of woodland walking | Single-group design |
| Patel_2013 | The long-term effects of a primary care physical activity intervention on mental health in low-active, community-dwelling older adults | Not a nature intervention |
| Heliker_2000 | The meaning of gardening and the effects on perceived well being of a gardening project on diverse populations of elders. | Qualitative study |
| Peacock_2005 | The mental and physical health outcomes of green exercise | School programmes with outdoor play, no clear nature focus |
| Miller_2018 | The Minne-Loppet Motivation Study: An Intervention to Increase Motivation for Outdoor Winter Physical Activity in Ethnically and Racially Diverse Elementary Schools | School programmes with outdoor play, no clear nature focus |
| Palmer_2020 | The Motor skills At Playtime intervention improves children's locomotor skills: A feasibility study. | School programmes with outdoor play, no clear nature focus |
| Palsdottir_2020 | THE NATURE STROKE STUDY; NASTRU: A RANDOMIZED CONTROLLED TRIAL OF NATURE-BASED POST-STROKE FATIGUE REHABILITATION. | Duplicate / already included |
| Turner-Stokes_2014 | The Northwick Park Therapy Dependency Assessment (NPTDA) scale: A psychometric analysis from a large multicentre dataset | Not an interventional study |
| Uijtdewilligen_2019 | The Park prescription study: development of a community-based physical activity intervention for a multi-ethnic Asian population | Qualitative study |
| Sarmiento_2017 | The recreovia of bogota, a community-based physical activity program to promote physical activity among women: Baseline results of the natural experiment al ritmo de las comunidades | Inappropriate comparison |
| Niedermeier_2019 | The role of anthropogenic elements in the environment for affective states and cortisol concentration in mountain hiking-a crossover trial | Not a behavioural change intervention |
| Mao_2016 | The salutary influence of forest bathing on elderly patients with chronic heart failure | Duplicate / already included |
| Demeyer_2017 | The survival effect of physical activity in patients with COPD: every step counts | Wrong publication type |
| Hoegmark_2020 | The wildman programme. A nature-based rehabilitation programme enhancing quality of life for men on long-term sick leave: Study protocol for a matched controlled study in Denmark | Ongoing study |
| Tse_2010 | Therapeutic effects of an indoor gardening programme for older people living in nursing homes | Not a nature intervention |
| Tse_2010 | Therapeutic effects of an indoor gardening programme for older people living in nursing homes. | Duplicate / already included |
| Connell_2007 | Therapeutic effects of an outdoor activity program on nursing home residents with dementia | Inappropriate comparison |
| NCT04656158_2020 | Therapeutic Effects of Horticulture on Anterior Cingulate Cortex Activation in People With Chronic Low Back Pain | Ongoing study |
| Gonzalez_2009 | Therapeutic horticulture in clinical depression: a prospective study. | Single-group design |
| Paddon_2020 | Therapeutic or detrimental mobilities? Walking groups for older adults | Not an interventional study |
| NCT04914572_2021 | Therapeutic Rainforest Study in Generating Positive Energy Among Undergraduate Students | Ongoing study |
| Bruijns_2021 | Training may enhance early childhood educators' self-efficacy to lead physical activity in childcare. | Not a nature intervention |
| Nost_2018 | Twelve-month effect of chronic pain self-management intervention delivered in an easily accessible primary healthcare service - a randomised controlled trial | Not a nature intervention |
| Williams_2011 | Type 2 diabetes and obesity: How effective is a lifestyle intervention at promoting physical activity and nutrition. | Wrong publication type |
| NCT02976506_2016 | Unsupervised Physical Activity in Elderly | Protocol without published results |
| Peacock_2021 | Use of Outdoor Education to Increase Physical Activity and Science Learning among Low-Income Children from Urban Schools | School programmes with outdoor play, no clear nature focus |
| Read_2011 | Using a goal setting list to help deliver an intervention in a randomised controlled trial | Not a nature intervention |
| NCT02150148_2014 | Vegetable Garden Feasibility Trial to Promote Function in Older Cancer Survivors | Duplicate / already included |
| NCT03997344_2019 | Veterans Nature Therapy (Vet Hike) | Protocol without published results |
| Kobayashi_2008 | Visiting a forest, but not a city, increases human natural killer activity and expression of anti-cancer proteins | Single-group design |
| Nowak_2020 | Vitamin D and indices of bone and carbohydrate metabolism in postmenopausal women subjected to a 12-week aerobic training program-The pilot study | Not a nature intervention |
| O'Brien_2011 | Volunteering in nature as a way of enabling people to reintegrate into society | Qualitative study |
| NCT03442998_2017 | Walking Green: developing an Evidence-base for Nature Prescriptions | Duplicate / already included |
| RBR-3h5394_2018 | Walking program for elderly women with high blood pressure | Not a nature intervention |
| Sui_2013 | Walking to limit gestational weight gain and keep fit during pregnancy - findings from the walk randomised trial. | Wrong publication type |
| Zhu_2021 | Waterfall Forest Environment Regulates Chronic Stress via the NOX4/ROS/NF-kappaB Signaling Pathway. | Animal study |
| N/A | What is preferable: treadmill or walking outdoors in order to improve walking ability after stroke? | Duplicate / already included |
| Coffey_2016 | When Pediatric Primary Care Providers Prescribe Nature Engagement at a State Park, Do Children â€œFillâ€ the Prescription? | Single-group design |
| Hillier_2020 | WHYoutdoors-Nature-based cancer care course | Wrong publication type |
| Irvine_2004 | Work breaks and well-being: The effect of nature on hospital nurses. | Wrong publication type |
| Swinburn_1998 | Written and verbal advice from general practitioners led to an increase in self-reported physical activity for sedentary patients | Not a nature intervention |
| NCT02400554_2015 | Yappalli - The Road to Choctaw Health | Not a nature intervention |
| Lewis_2009 | Youth and nature: Assessing the impact of an integrated wellness curriculum on nature based play and nature appreciation for youth in out-of-school time recreation programming. | Wrong publication type |
