## Supplementary File S4: Summary of intervention characteristics for "Nature prescriptions: a scoping review with a nested meta-analysis"

| Study ID | Nature setting | Type of activities promoted | Indoor or outdoor? | Referring institution | Social Cognitive Framework | | | Co-intervention |
| --- | --- | --- | --- | --- | --- | --- | --- | --- |
|  |  |  |  |  | **Cognitive factors** | **Behavioural factors** | **Environmental factors** |  |
| Akgoz_2020 | Parks | Walking | Outdoor | Family health centre | Y | Y | Y | - |
| Ameli_2021 | Public gardens | Walking | Outdoor | Hospital | N | Y | Y | - |
| Arbillaga-Etxarri_2017 | Parks & beaches | Walking | Outdoor | Health provider | Y | Y | Y | - |
| Baba_2021 | Parks | Cleaning; Gardening | Outdoor | Care facility | N | Y | Y | - |
| Ballew_2018 | Arboretum | Relaxation | Outdoor | - | N | Y | Y | - |
| Bang_2017 | Forests | Walking | Outdoor | - | Y | Y | Y | Stress & depression management lessons |
| Bang_2018 | Forests | Camping; Sports; Group Games | Outdoor | Welfare centre | Y | Y | Y | - |
| Barton_2012 | Parks, countryside areas & nature reserves | Walking | Outdoor | Community service centre | N | Y | Y | - |
| Barton_2015 | School fields & nearby green spaces | Orienteering Activities | Outdoor | School | N | Y | Y | - |
| Bielinis_2021 | Forests | Relaxation | Outdoor | - | N | Y | Y | - |
| Bloom_2017 | Parks | Walking | Outdoor | Employer | N | Y | Y | - |
| Brito_2020 | Arboretum | Walking | Outdoor | - | N | Y | N | - |
| Brown_2014 | Streetscape trees & grass | Walking | Outdoor | Employer | Y | Y | Y | - |
| Calogiuri_2016 | Forests, grass-yard | Biking; Strength Training | Outdoor | Employer | Y | Y | Y | - |
| Chun_2017 | Forests | Walking; Meditation | Outdoor | Welfare centre | N | Y | Y | - |
| Cimprich_2003 | Community gardens | Bird-Watching; Relaxation; Walking | Outdoor | Hospital | N | Y | Y | - |
| Clutterbuck_2020 | Parks | Sports | Outdoor | Community service centre | N | Y | Y | - |
| Cohen_2017 | Parks | Fitness Classes | Outdoor | - | Y | Y | Y | - |
| Corazon_2018 | Arboretum | Mindfulness; Relaxation; Walking | Outdoor | Health service centre | N | Y | Y | - |
| Demark-Wahnefried_2018 | Home gardens | Gardening | Indoor | Hospital | Y | Y | Y | - |
| Detweiler_2015 | Institutional gardens | Gardening | Indoor | Hospital | N | Y | Y | - |
| Djernis_2021 | Therapy gardens | Yoga; Walking; Mindfulness | Outdoor | - | Y | Y | Y | - |
| Elsey_2018 | Farms | Farming | Outdoor | Probation service centre | N | Y | N | - |
| Finkelstein_2013 | Nature reserves & parks | Hiking | Outdoor | - | Y | Y | Y | - |
| Flowers_2018 | Parks | Cycling | Outdoor | - | N | Y | N | - |
| Fruhauf_2016 | Greenspace | Walking | Outdoor | Hospital | N | Y | Y | - |
| Garshol_2020 | Farms | Farming | Outdoor | Day care service centre | N | Y | N | - |
| Gascon_2020 | Community gardens | Gardening | Outdoor | Community garden centre | N | Y | Y | - |
| Gladwell_2016 | Nature trails | Walking | Outdoor | - | N | Y | N | - |
| Grazuleviciene_2015 | Parks | Walking | Outdoor | Clinic/hospital | N | Y | Y | - |
| Han_2016 | Forests | Relaxation; Music Therapy | Outdoor | Employer | N | Y | Y | - |
| Han_2018 | Farms | Gardening | Outdoor | Health service centre | N | Y | Y | - |
| Heilmayr_2018 | Community gardens & other urban green spaces | Gardening; Relaxation | Indoor & outdoor | - | N | Y | N | - |
| Hoffman_2018 | Parks | Structured Games & Sports | Outdoor | Clinic/hospital | N | Y | Y | Swimming lessons, peer support, cooking classes, clinic treatment |
| Horiuchi_2015 | Forests | Relaxation; Walking | Outdoor | Local authority | N | Y | Y | - |
| Jeon_2021 | Forests | Relaxation; Walking | Outdoor | Probation service centre | N | Y | Y | - |
| Kam_2010 | Institutional gardens | Gardening | Outdoor | Rehabilitation centre | N | Y | Y | Group support |
| Kang_2021 | Forests | Art; Relaxation | Outdoor | Counselling centre | N | Y | Y | - |
| Kim_2018a | Institutional gardens | Gardening; Craft | Indoor & outdoor | Community service centre | N | Y | Y | - |
| Kim_2018b | Parks | Resistance Exercises | Outdoor | Welfare centre | N | Y | N | - |
| Kim_2021 | Forests | Stretching; Walking; Exercises; Meditation | Outdoor | - | N | Y | Y | - |
| Kobayashi_2018 | Forests | Walking | Outdoor | - | N | Y | N | - |
| Koselka_2019 | Forests | Walking | Outdoor | - | Y | Y | Y | - |
| Lacharite-Lemieux_2015 | Parks | Exercises | Outdoor | - | N | Y | N | - |
| South_2021 | Parks, vacant lots & schoolyards | Reading; Walking; Gardening; Music | Outdoor | Clinic/hospital | Y | Y | Y | - |
| Lee_2014 | Forests | Walking | Outdoor | Clinic/hospital | N | Y | Y | - |
| Leiros-Rodriguez_2014 | Parks | Balance Exercises | Outdoor | Senior centre | N | Y | Y | - |
| Li_2016 | Forests | Walking | Outdoor | - | N | Y | N | - |
| Liu_2020 | Parks | Exercises | Outdoor | - | N | Y | Y | - |
| Makizako_2020 | Community gardens | Gardening | Outdoor | - | N | Y | Y | - |
| Mao_2012a | Forests | Walking | Outdoor | - | N | Y | Y | - |
| Mao_2012b | Forests | Walking | Outdoor | - | N | Y | Y | - |
| Mao_2017 | Forests | Walking | Outdoor | - | N | Y | Y | - |
| McEwan_2019 | Urban green spaces | Any | Outdoor | - | Y | Y | Y | - |
| Miller_2020 | Parks | Exercises | Outdoor | Clinic/hospital | N | Y | N | - |
| Mohamed_2018 | Community gardens | Gardening | Indoor & outdoor | - | Y | Y | Y | Exercise programmes |
| Morris_2021 | Any | Any | Outdoor | Clinic/hospital | Y | Y | Y | - |
| Muller-Riemenschneider_2020 | Parks | Any | Outdoor | Community service centre | Y | Y | Y | - |
| Ng_2018 | Community gardens, parks, nature reserves | Gardening; Stretching; Walking | Indoor & outdoor | - | N | Y | Y | - |
| Ngo_2014 | Nature reserves & parks | Hiking; Sports | Outdoor | - | Y | Y | Y | Step prescription |
| Oh_2018 | Farms | Gardening | Outdoor | Clinic/hospital; Mental rehabilitation centre | Y | Y | Y | - |
| Palsdottir_2020 | Gardens | Walking; Cycling; Exercises; Gardening; Relaxation; Socialising | Indoor & outdoor | Clinic/hospital | N | Y | Y | - |
| Park_2010 | Forests | Walking; Relaxation | Outdoor | - | N | Y | Y | - |
| Park_2020a | Forests | Meditation; Walking; Massage | Outdoor | - | N | Y | Y | - |
| Park_2020b | Community gardens | Gardening | Outdoor | Senior centre | N | Y | Y | - |
| Payne_2020 | Any | Any | Outdoor | - | Y | Y | Y | - |
| Plotnikoff_2017 | Parks | Strength & Aerobic Exercises | Outdoor | Health service centre | Y | Y | Y | - |
| Razani_2018 | Parks | Any | Outdoor | Clinic/hospital | Y | Y | Y | - |
| Ryu_2020 | Any green space | Cycling | Outdoor | Clinic/hospital; Mental health centre | Y | Y | Y | - |
| Sales_2017 | Parks | Exercises | Outdoor | - | N | Y | Y | - |
| Serrat_2020 | Any | Hiking; Yoga; Nordic Walking; Photography; Relaxation | Outdoor | Clinic/hospital | N | Y | Y | Pain neuroscience education, cognitive behavioral therapy, mindfulness training |
| Shin_1999 | Parks | Walking | Outdoor | Senior centre | N | Y | Y | - |
| Siu_2020 | Community gardens | Gardening | Outdoor | Vocational rehabilitation centre | N | Y | N | - |
| Song_2013 | Forests | Relaxation | Outdoor | - | N | Y | N | - |
| Song_2019 | Forests | Walking | Outdoor | - | N | Y | Y | - |
| Stigsdotter_2018 | Therapy gardens | Mindfulness; Relaxation; Walking | Outdoor | Health service centre; insurance company; clinic/hospital | N | Y | Y | - |
| Sung_2012 | Forests | Relaxation | Outdoor | Health centre | Y | Y | Y | Cognitive-behavioural education |
| Takayama_2014 | Forests | Walking; Relaxation | Outdoor | - | N | Y | N | - |
| Tharrey_2020 | Commuity gardens | Gardening | Outdoor | Community garden centre | N | Y | Y | - |
| Turner_2017 | Woodlands | Running | Outdoor | - | N | Y | N | - |
| Ura_2021 | Rice fields | Farming | Outdoor | Clinic/hospital | N | Y | Y | - |
| VanDenBerg_2011a | Community gardens | Gardening | Outdoor | Allotment complex | N | Y | Y | - |
| VanDenBerg_2011b | Woodlands | Group Activities | Outdoor | Care farm | N | Y | Y | - |
| Verra_2012 | Greenhouses | Gardening | Indoor | Clinic/hospital | Y | Y | Y | - |
| Vujcic_2017 | Botanical gardens | Gardening; Mindfulness; Sunbathing; Meditation; Art Therapy; Socialising | Outdoor | Clinic/hospital | N | Y | Y | - |
| Wang_2018 | Forests | Walking | Outdoor | - | N | Y | Y | - |
| Wexler_2021 | Parks | Any physical activity | Outdoor | - | N | Y | Y | - |
| Wichrowski_2005 | Greenhouses & institutional gardens | Gardening | Indoor | Clinic/hospital | Y | Y | Y | - |
| Willert_2014 | Arboretum, forests & beaches | Socialising; Relaxation; Exercises; Yoga; Gardening; Walking | Outdoor | Job rehabilitation centre | N | Y | Y | Job counselling |
| Wong_2021 | Community gardens, home gardens & parks | Gardening; Walking | Indoor & outdoor | - | N | Y | Y | - |
| Wu_2020 | Forests | Walking | Outdoor | - | N | Y | Y | - |
| Yi_2021 | Forests | Qigong; Walking | Outdoor | Health service centre; Senior centre; Culture centre | N | Y | Y | - |
| Zhu_2016 | Community gardens | Gardening | Indoor & outdoor | Clinic/hospital | Y | Y | Y | - |
