## Supplementary figures and images for "Nature prescriptions: a scoping review with a nested meta-analysis"

### Supplementary Figure S1: Distribution by year

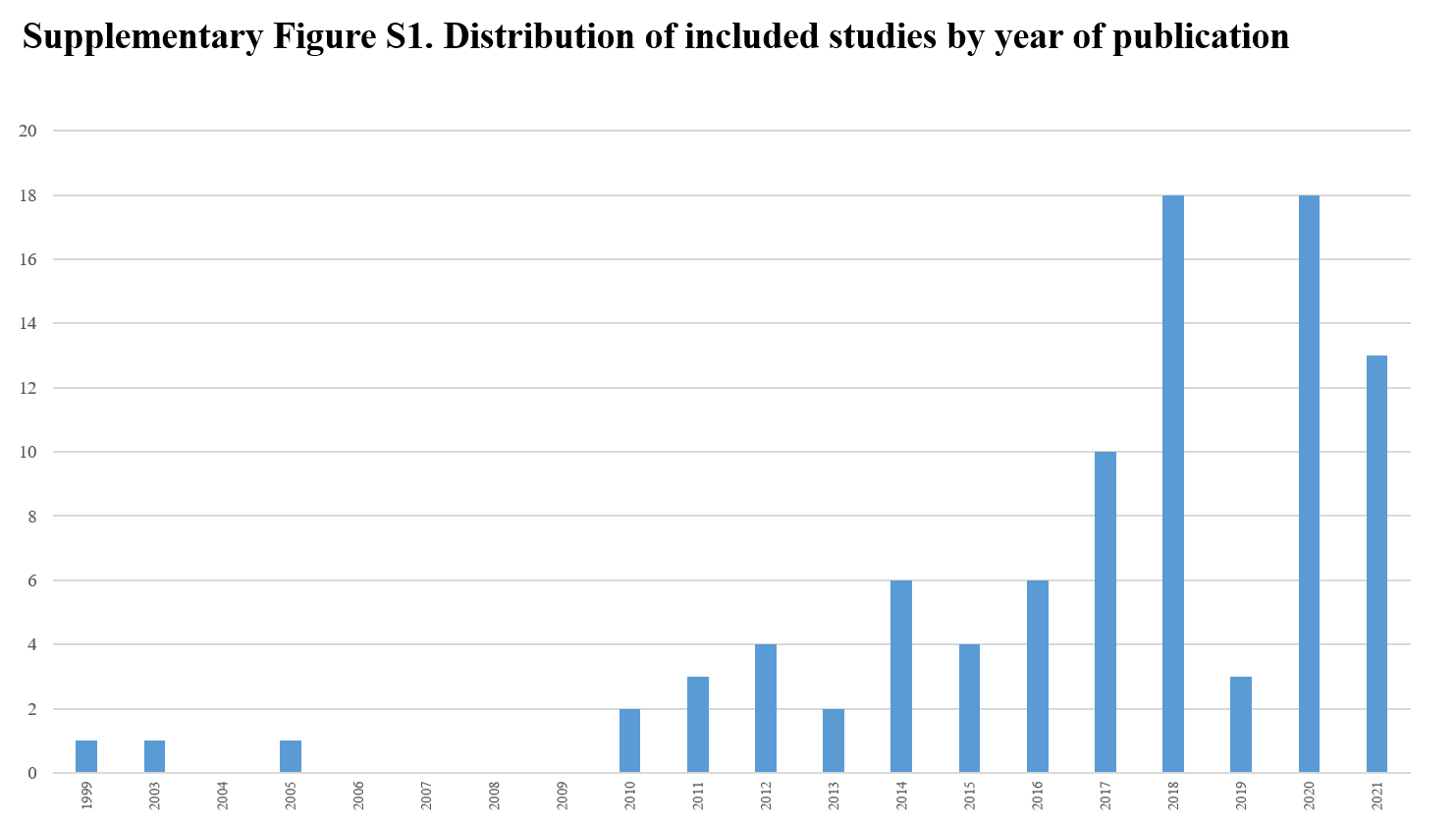

### Supplementary Figure S2: Distribution by location

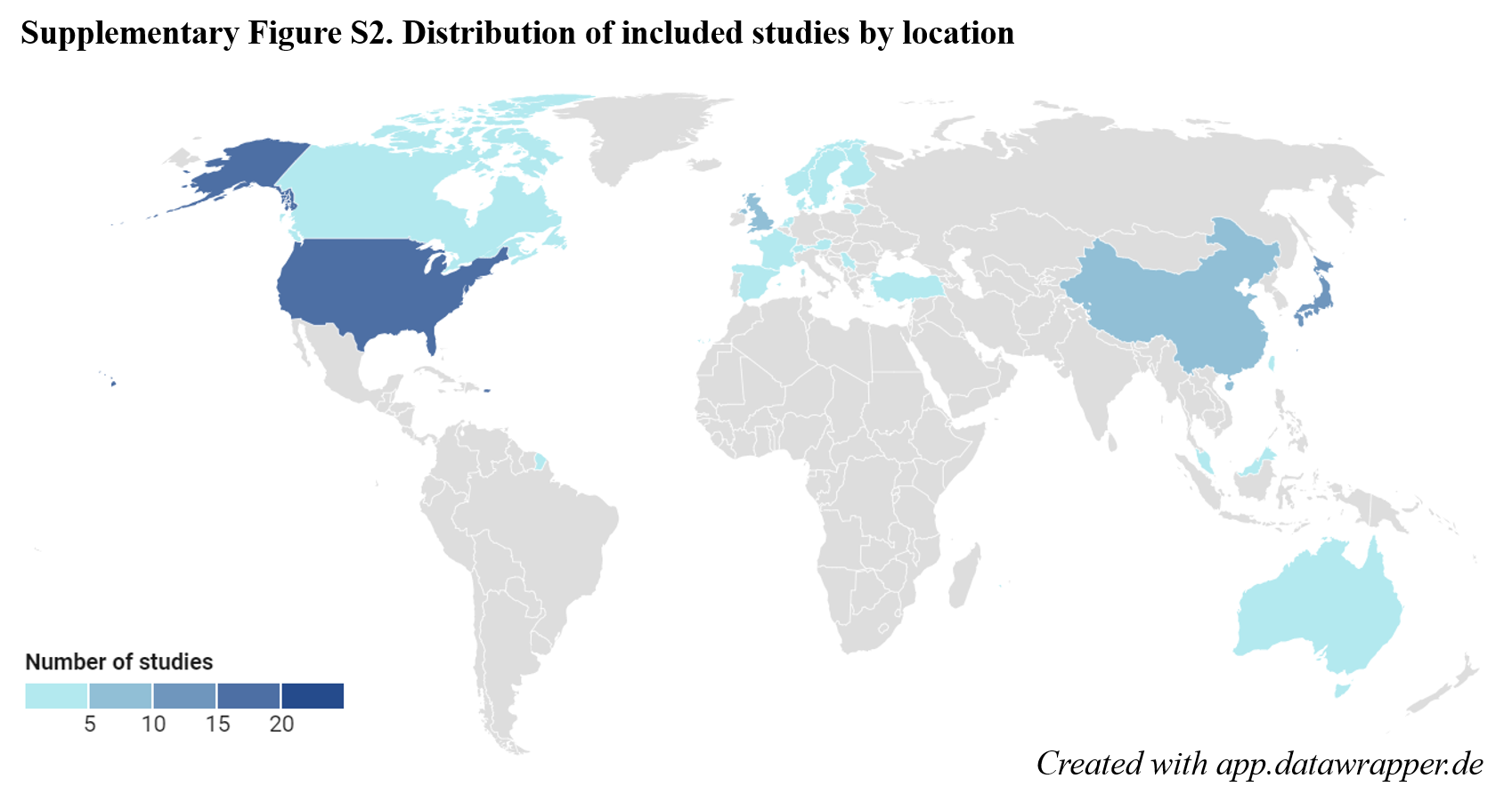
